# Linoleoyl-lysophosphatidylcholine suppresses immune-related adverse events due to immune checkpoint blockade

**DOI:** 10.1101/2024.08.07.24310974

**Authors:** Ian T. Mathews, Priyanka Saminathan, Mir Henglin, Mingyue Liu, Namratha Nadig, Camille Fang, Kysha Mercader, Serena J. Chee, Allison M. Campbell, Abhijit A. Patel, Saumya Tiwari, Jeramie D. Watrous, Karthik Ramesh, Martina Dicker, Khoi Dao, Melissa A. Meyer, Pekka Jousilahti, Aki S. Havulinna, Teemu Niiranen, Veikko Salomaa, Leo A.B. Joosten, Mihai G. Netea, Pan Zheng, Mitchell Kronenberg, Sandip Pravin Patel, J. Silvio Gutkind, Christian Ottensmeier, Tao Long, Susan M. Kaech, Catherine C. Hedrick, Susan Cheng, Mohit Jain, Sonia Sharma

## Abstract

Immune related adverse events (irAEs) after immune checkpoint blockade (ICB) therapy occur in a significant proportion of cancer patients. To date, the circulating mediators of ICB-irAEs remain poorly understood. Using non-targeted mass spectrometry, here we identify the circulating bio-active lipid linoleoyl-lysophosphatidylcholine (LPC 18:2) as a modulator of ICB-irAEs. In three independent human studies of ICB treatment for solid tumor, loss of circulating LPC 18:2 preceded the development of severe irAEs across multiple organ systems. In both healthy humans and severe ICB-irAE patients, low LPC 18:2 was found to correlate with high blood neutrophilia. Reduced LPC 18:2 biosynthesis was confirmed in preclinical ICB-irAE models, and LPC 18:2 supplementation *in vivo* suppressed neutrophilia and tissue inflammation without impacting ICB anti-tumor response. Results indicate that circulating LPC 18:2 suppresses human ICB-irAEs, and LPC 18:2 supplementation may improve ICB outcomes by preventing severe inflammation while maintaining anti-tumor immunity.

## Introduction

ICB therapies targeting PD-1/PD-L1 or CTLA-4 signaling have transformed solid tumor immune-oncology (I/O) and are indicated for more than 20 cancer types, with greater than 40% of cancer patients in the US currently eligible for ICB therapy^1^. Improving ICB outcomes is therefore of high clinical value and subject to intense investigation. ICB-driven tumor regression is positively influenced by an abundance of tumor neoantigens and high expression of certain immune markers^2^. However, ICB therapy, is often associated with the development of treatment-associated systemic inflammation and subsequent adverse events (ICB-irAE). Severe grade III/IV ICB-irAEs in particular drive critical morbidity in a significant fraction of cancer patients. Unlike other drug-induced hypersensitivities, such as toxic epidermal necrosis (TEN)/Stevens-Johnson Syndrome in HLA-B*15:02 carriers receiving carbamazepine, no definitive gene loci have been associated with severe ICB-irAEs to guide reliable risk-benefit assessments. The potential for co-morbidity is unlikely to guide clinical decision making in offering ICB, as indications for cancer immunotherapy are almost exclusively prescribed for advanced and treatment refractory malignancies. Thus, effective treatments for mitigating ICB-irAEs without interfering with the efficacy of anti-tumor response are a significant unmet need in immune oncology today.

ICB-irAEs manifest most often with the CTLA-4 blocker ipilimumab^1^, and in combination therapy with the PD-1 blocker nivolimumab severe irAEs have been described in up to 60% of individuals^3^, with ∼40% of cases resulting in treatment discontinuation^4^ and ∼1 percent in death^5^. Current clinical guidelines for management of ICB-irAEs^6^ recommend administration of corticosteroids and other broadly-acting immunosuppressive agents after the onset of ICB-irAE. In this study we used non-targeted mass spectrometry analysis of plasma from cancer patients undergoing ICB therapy for solid tumor. We uncovered a circulating factor, the bio-active signaling lipid LPC 18:2, whose post-therapy loss preceded the subsequent development of severe irAEs across several different organ systems and increased blood neutrophilia. Functional studies in preclinical models of ICB-irAE showed that LPC 18:2 metabolism is dysregulated in relation to inflammation, and LPC 18:2 supplementation reduced irAE severity and lowered circulating pro-inflammatory neutrophils *in vivo*. Together, these studies link bio-active lipids with irAE severity, and suggest that modulating LPC 18:2 concentrations in the blood may suppress severe irAEs.

## Results

### Circulating lysolipids are depleted after development of ICB-irAEs and autoimmune disease

ICB-irAEs affect multiple organ systems, suggesting that systemic factors might influence their severity. We and others have found that circulating small molecule metabolites modulate both oncogenic and immunological phenotypes in health and disease^7^. Thousands of bio-active lipids with signaling function control cellular states of inflammation and immunity^8^, including oxylipins, bile acids and related molecules with established immune-modulatory functions across multiple organs, including colon, lung, skin, heart and vascular systems^9–12^. To uncover previously unknown molecules regulating ICB-irAEs, we assayed thousands of circulating bio-active lipids across three independent human studies of anti-CTLA-4 or anti-PD-1 therapy for solid tumor, totaling over 150 patients and 750 plasma bio-samples (**Supplementary Table 1**). In the discovery cohort, corresponding to patients with advanced melanoma treated with 3mg/kg ipilimumab (cohort 1, n = 65, **Supplementary Table 2**)^13^, serial plasma bio-samples were first analyzed using non-targeted liquid chromatography mass spectrometry (LC-MS)^14^. Nearly 6000 distinct small molecule bio-active lipid metabolites were measured in human plasma, with significant loss in a subset of metabolites over time observed in association with the development of ipilimumab-associated irAEs (**Fig 1a**). Chemical network analysis of irAE-associated metabolites revealed them to be most similar to lysophosphatidylcholines (LPC) (**Extended Data Fig 1a**). Structural elucidation was confirmed using synthesized standards, and metabolites were identified as palmitoyl-lysophosphatidylcholine (LPC (16:0)) and linoleoyl-lysophosphatidylcholine (LPC 18:2) (**Fig 1b**), monoacyl metabolites of membrane phosphocholines (PC)^15^. LPCs comprise a superfamily of bio-active signaling lipids that are broadly implicated in regulation of immunity and inflammation via cell-intrinsic effects on a number of different immune cells in the blood, including neutrophils, monocytes/macrophages and lymphocyte populations^16^. Importantly, ICB-irAE associations were specific to LPC (16:0) and LPC 18:2, and were not observed for an additional 30 LPC metabolites or for lysophosphatidylethanolamines (LPE), lysophosphatidylinositols (LPI), or lysophosphatidic acid (LPA) metabolites (**Supplementary Table 3**). Temporal analysis at baseline through twelve weeks post-ipilimumab treatment revealed a drop in LPC 18:2 and LPC (16:0) concentrations as early as three weeks following ICB treatment, specifically in patients who developed severe grade III/IV irAEs but not mild grade I/II irAEs (**Fig 1c, Extended Data Fig 1b**). Baseline, pre-therapy LPC levels were not associated with ICB-irAEs (**Extended Data Fig 1c**). Furthermore, baseline or post-therapy changes in LPC 18:2 and LPC (16:0) were not significantly associated with anti-tumor immunity by RECIST 1.1 metrics^17^ (**Supplementary Table 3**).

**Figure 1.**
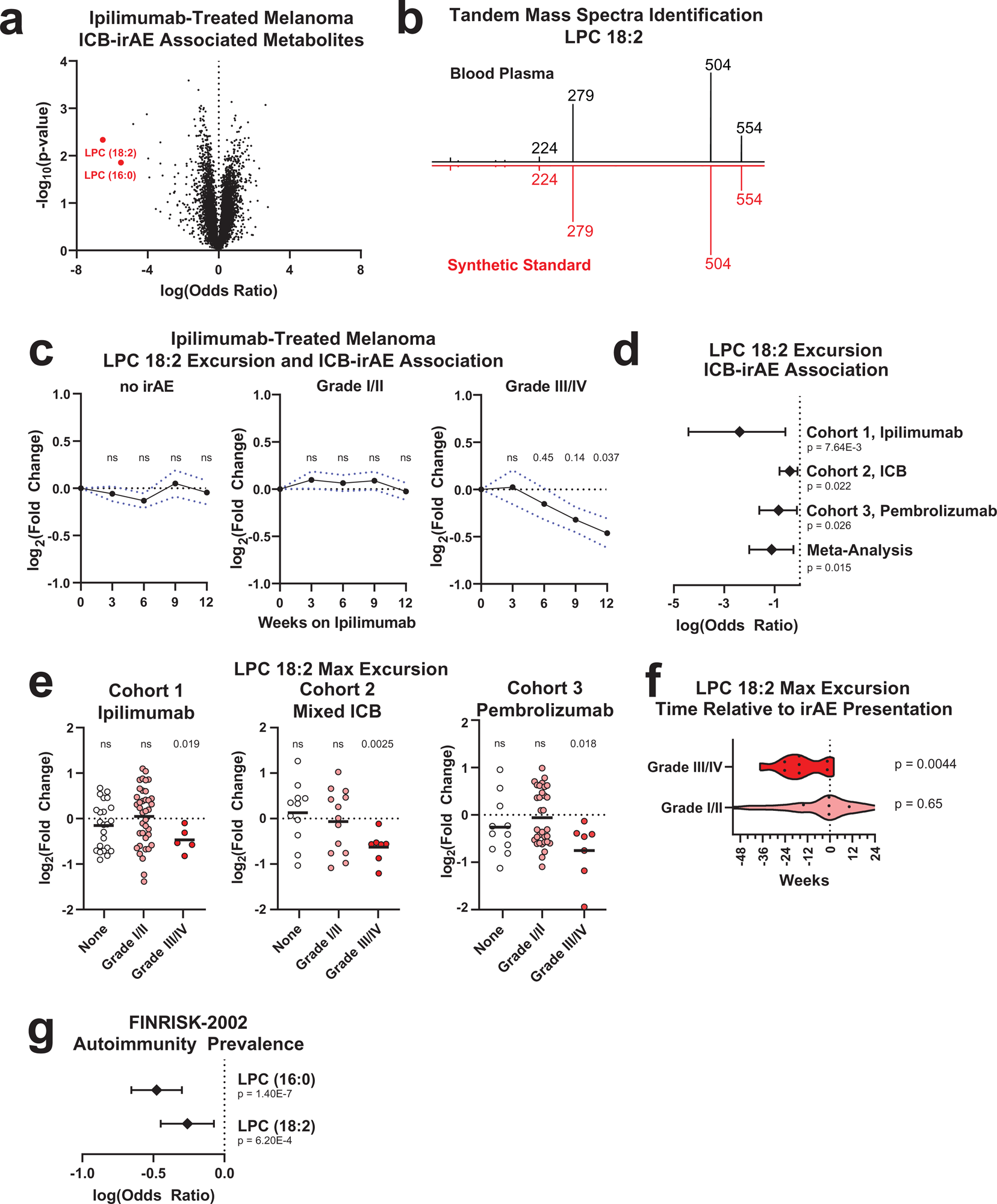
Identification of circulating lysolipids depleted after development of ICB irAEs and autoimmunity. (**a**) Volcano plot of irAE severity logistic regressions in ipilimumab-treated melanoma patients (cohort 1, n = 65), denoting maximum excursion (change) from baseline samples of 5951 ubiquitous, bioactive lipid metabolites. Metabolites corresponding to LPC 18:2 and LPC (16:0) (red) are highlighted. Odds ratio normalized to a natural logarithm. (**b**) Tandem mass spectra of *m/z* 554.3019, corresponding to LPC 18:2 [M+Cl^-^]. (**c**) Mean excursion of LPC 18:2 in cohort 1, error = SEM. (**d**) Forest plots of natural log odds ratio of irAE severity for LPC 18:2. Error = 95% CI. (**e**) Maximum excursion of LPC 18:2 in three cohorts of ICB-treated patients, bars = mean. (**f**) Time of maximum LPC 18:2 excursion from baseline sampling relative to reported irAE date in cohort 3. (**g**) Natural log odds ratio of prevalent autoimmune pathologies in FINRISK-2002 (case: n = 158, control: n = 1712). Error = 95% CI. Logistic regression with patient age, patient sex as covariates (Fig 1a**, 1d, 1g**); one-sided Student’s T-test (Fig 1c**, 1e-f**).

Clinical and preclinical evidence support both overlapping and distinct mechanisms of action for anti-CTLA-4 and anti-PD-1 blockers in solid tumor I/O^18^. To explore the generalizability of LPC associations with severe ICB-irAEs, we examined two additional patient cohorts (**Supplementary Table 1**). In a registry study of cancer patients with solid tumor, either treated with ipilimumab and nivolumab or with nivolumab or pembrolizumab alone (cohort 2, n = 32), significant reduction in LPC 18:2 and LPC (16:0) concentrations were confirmed in patients who developed severe grade III/IV irAEs (**Fig 1d-e, Extended Data Fig 2a-b**). In a prospective Phase I trial of pembrolizumab for advanced non-small cell lung cancer (NSCLC) or melanoma^19^ (cohort 3, n= 53), significant reduction of plasma LPC 18:2 and LPC (16:0) was also observed only in cases of severe irAEs (**Fig 1d-e**). Despite differences across tumor types and ICB regimens, the degree of association between LPCs and severe irAEs was comparable among the three independent human studies (**Fig 1d**). Loss of plasma LPC 18:2 was observed in relation to irAEs occurring in different organ systems, including gastrointestinal irAEs and pulmonary irAEs (**Extended Data 2c**), suggesting a broadly applicable mechanism. Moreover, the reduction in circulating LPC 18:2 preceded the onset of severe irAEs by a mean of 14.9 weeks (**Fig 1f**). Together, these data show that unique lysolipids, LPC (16:0) and LPC 18:2, are selectively diminished in the blood of solid tumor cancer patients who received anti-CTLA-4 or anti-PD-1 ICB therapy, and subsequently predict the development of severe irAEs across diverse organ systems.

To determine whether the link between LPC and systemic inflammation is generalizable, we examined LPC (16:0) and LPC 18:2 concentrations in plasma samples from 158 community dwelling individuals with acquired autoimmune disease, including systemic lupus erythematosus (SLE), rheumatoid arthritis, and inflammatory bowel disease, as well as 1712 healthy controls from the FINRISK-2002 study (**Supplementary Table 4**)^20^. Participants with autoimmune diseases had lower circulating LPC 18:2 and LPC (16:0) concentrations (**Fig 1g**) relative to healthy counterparts, suggesting that similar to ICB-irAEs, autoimmune-mediated inflammation is also associated with reduced circulating LPC. Similar to other lysolipids, LPC 18:2 is derived from dietary fats and is specifically and most commonly produced from esterification of dietary linolenic acid and desaturation of dietary linoleic acid. To explore the relationship between dietary exposure and circulating LPC concentrations, dietary data from FINRISK-2002 was examined, confirming that intake of foods rich in fat significantly increased LPC 18:2 and LPC (16:0) concentrations in the blood (**Supplementary Table 5**). Food-specific determinants of significant change in blood LPC included a low-fat diabetic diet, consistent with the reported association of these lipids with prevalent diabetes mellitus^21^, as well as carbohydrate-rich and oil-rich foods. However, components of exclusionary or low-retention diets commonly adopted for chronic inflammatory colitis, including lactose, fiber, and gluten, did not associate significantly with changes in concentration of blood LPC 18:2.

### Lower LPC 18:2 concentrations correlate with higher neutrophil numbers in the blood of healthy humans and ICB-irAE patients

LPC concentrations were next measured in association with features of systemic immunity^22^. Across healthy individuals in the 500 Functional Genomics (500FG) community-dwelling cohort, for whom dense measures of circulating immune cells have been reported, circulating LPC 18:2 concentrations were specifically and inversely associated with neutrophil counts (**Fig 2a** and **Extended Data Fig 3a**). In contrast, LPC (16:0) concentrations were more broadly correlated with multiple inflammatory and non-inflammatory immune cell types in the blood, including several lymphocyte and monocyte populations (**Extended Data Fig 3a**). Similarly, LPC 18:2 was also inversely correlated to absolute neutrophil numbers in a disease-free subset of FINRISK-2002 (**Fig 2b** and **Supplementary Table 4**), although not with circulating molecules such as immunoglobulin (**Extended Data Fig 3b**). Importantly, high neutrophil-to-lymphocyte ratios after ICB therapy are an established risk factor for severe ICB-irAEs^23^. Accordingly, blood panels from irAE patients in cohort 1 revealed enhanced blood neutrophilia (**Fig 2c**), in contrast to lymphocyte and monocyte excursions which showed no significant association with irAEs (**Extended Data Fig 4a**). Corticosteroid therapy promotes blood neutrophilia^24^, and comparison of neutrophil counts in the steroid-naïve and steroid-treated ICB-irAE patients (**Fig. 2c**) revealed significant increases in both patient populations. In contrast, baseline differences in leukocyte populations were not observed between irAE severity groups (**Extended Data Fig 4b**). In cohorts 1 and 2, LPC 18:2 concentrations also inversely correlated with neutrophil numbers (**Fig 2d**). Together, results demonstrate that lower blood concentrations of LPC 18:2 are linked to elevated neutrophil numbers across diverse healthy individuals and ICB-irAE patients.

**Figure 2.**
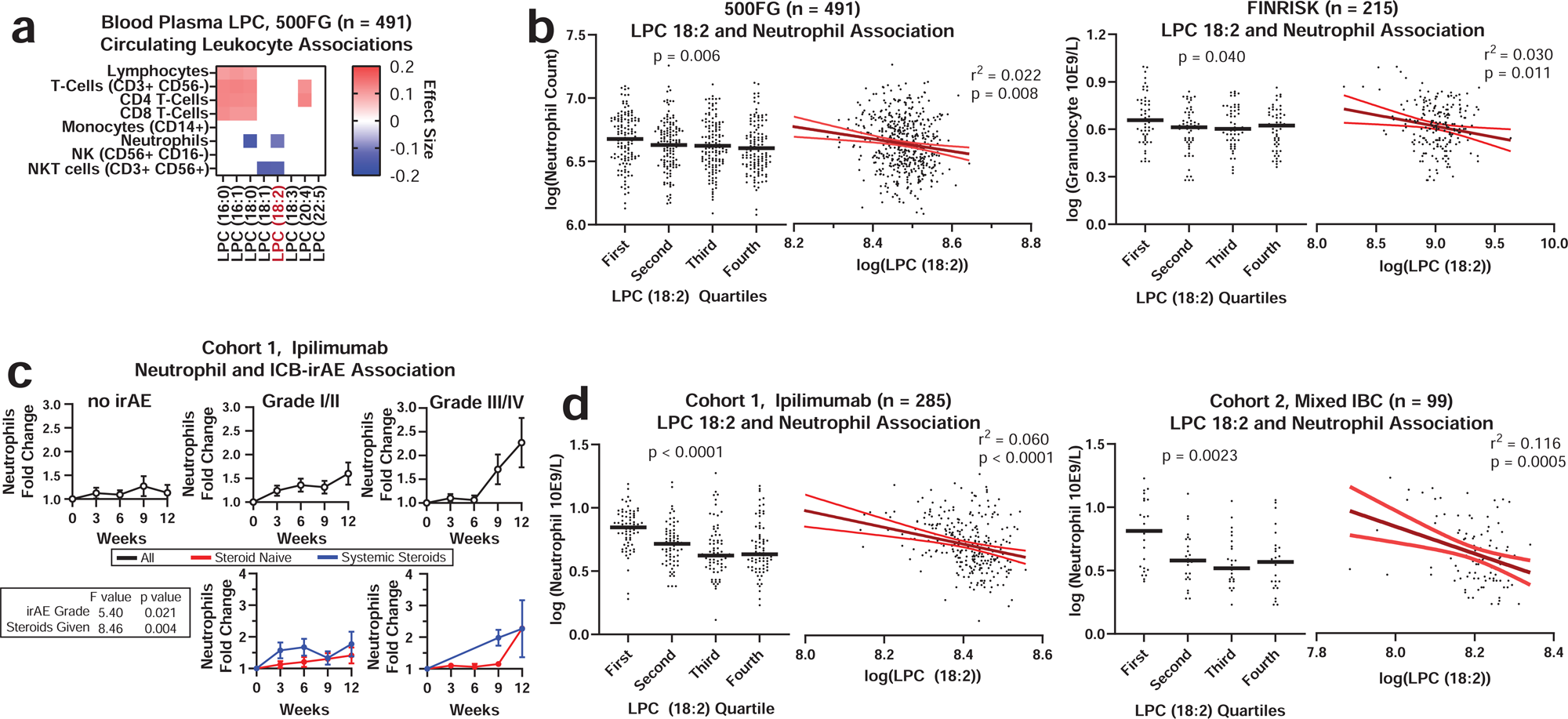
LPC 18:2 concentrations inversely correlate with neutrophil cellularity in the blood of healthy humans and ICB irAE patients. (**a**) Significant associations by linear regression between LPC 18:2 and lymphoid and myeloid populations in matched blood from healthy individuals (500FG, n = 491, filled color indicates p = 0.05 following Bonferroni multiple hypothesis correction, blue negative correlation, red positive). (**b**) Quartiles and scatter plots of LPC 18:2 concentration and neutrophil counts as measured by multi-color flow cytometry (500FG, left) or by blood analyzer (FINRISK-2002, right). Bars = mean, black line = linear regression, red lines = standard deviation. (**c**) Fold change of neutrophil counts relative to baseline sampling in cohort 1 ipilimumab treated melanoma patients, all (top) and stratified by corticosteroid administration (bottom). Error = SEM. (**d**) Quartiles and scatter plots of LPC 18:2 concentration and neutrophil counts in ICB cohort 1 and cohort 2. Bars = mean. Linear regression with patient age, patient sex as covariates (Fig 2a); One-way ANOVA with test for linear trend (Fig 2b**, 2d**); two-way ANOVA of linear model with patient age, patient sex as covariates (Fig 2c). Significance derived from linear regression including patient and timepoint as covariates to control for internal correlation.

### *De novo* LPC 18:2 biosynthesis is impaired in mouse models of ICB-irAEs and colitis

To study the functional relationship between changes in LPC and development of ICB *in vivo*, a mouse model of ICB-irAEs corresponding to human *CTLA-4* gene knock-in mice (*CTLA-4^h/h^*) was examined (**Fig 3a**, *top*). After ipilimumab treatment or ipilimumab and anti-mouse PD-1 combination treatment, *CTLA-4^h/h^* mice develop graded, multi-organ irAEs affecting the heart, liver, lung, kidney, salivary gland and colon during the same therapeutic window as the tumor regression response^25^. Mass spectrometry analysis demonstrated significant reduction of LPC 18:2 concentrations in the blood of *CTLA-4^h/h^* mice after ICB treatment (**Fig 3b**, *left*), which inversely correlated with the differences in irAE severity observed after single or combination ICB treatment (**Fig 3b**, *right*). Relative to LPC 18:2, reduction of LPC (16:0) was less pronounced in the ipilimumab-treated mice, and was not significantly associated with irAE severity (**Extended Data Fig 5a**). Together, these data show that loss of circulating LPC 18:2 after ICB treatment is conserved in preclinical mouse models of ICB-irAEs.

**Figure 3.**
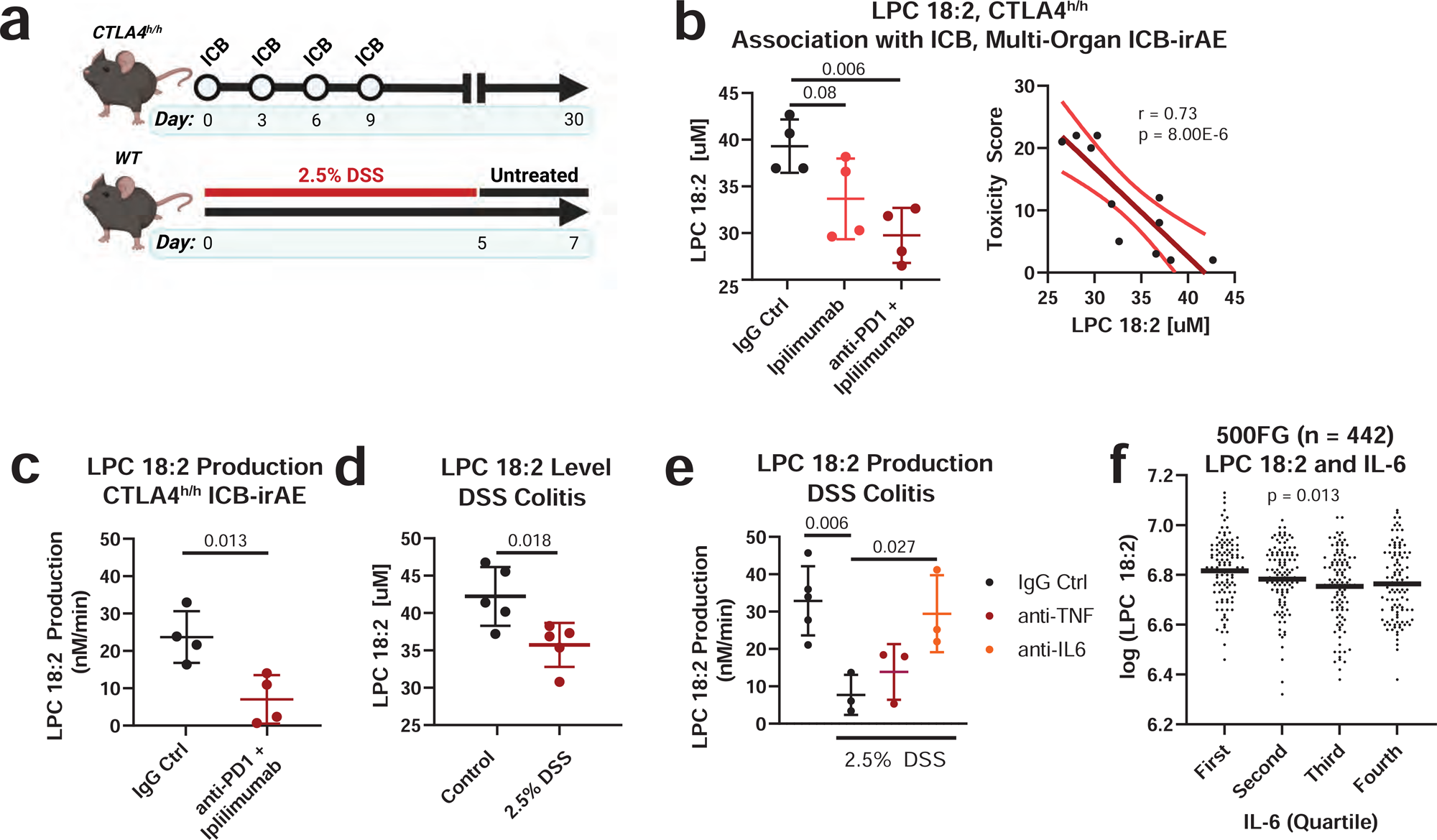
LPC 18:2 biosynthesis is impaired in mouse models of ICB irAEs and colitis. (**a**) Diagram of experimental design for *CTLA-4*^h/h^ model of ICB-irAE and DSS colitis. (**b**) LPC 18:2 concentration in terminal blood of CTLA-4^h/h^ C57BL/6 mice treated with 100 ug anti-human IgG1 isotype control (hIgFc), 100 ug ipilimumab (Ipi), or 100 ug ipilimumab + 100 ug anti-mouse PD-1 monoclonal antibody (Ipi + anti-PD-1). n = 4 per group. Error = SD. Bar = mean. Right: correlation plot of LPC 18:2 concentration to pathology score. Linear regression in dark red, 95% confidence interval in light red. (**c**) LPC 18:2 rate of production from blood plasma on Day 28 after ipilimumab + anti-PD-1 therapy *CTLA-4^h/h^* mice. n = 4 per group. Error = SD. Bar = mean. (**d**) LPC 18:2 concentration in terminal blood of 2.5% DSS-treated C57BL/6. n = 5 per group. Error = SD. Bar = mean. (**e**) *De novo* LPC 18:2 production in terminal blood of 2.5% DSS mice treated with 100ug control IgG, 100ug anti-TNF, or 100ug anti-IL-6 for 6 hours. n = 3-5 per group, Error = SD. Bar = mean. (**f**) Quartiles of IL-6 cytokine (pg/mL) and LPC 18:2 concentration in 500FG. Bars = mean. Student’s T-test (Fig. 3c, Fig. 3d), one-way ANOVA with Dunnett’s multiple comparisons test (Fig. 3b, Fig. 3e, Fig 3f).

Hydrolysis of membrane PC to produce LPC and subsequent conversion of LPC back to PC occurs via the Lands cycle^26^, a homeostatic process for maintenance of the diversity of membrane fatty acid composition in cells and tissues. Interestingly, evidence suggests that Lands cycle activity is dynamically regulated during inflammation through regulated expression of phospholipases (PLA) in inflamed extracellular spaces^27^, as observed for elevated soluble PLA2-IIA concentrations in the synovial fluid of rheumatoid arthritis patients^28^ and in the serum of severe COVID-19 patients^29^. To assess changes in PC hydrolysis as a possible mechanism for the loss of LPC 18:2 in ICB-irAE mice, conversion of exogenous 18:2 (*cis*) PC (1,2-dilinoleoyl-sn-glycero-3-phosphocholine or DLPC) to LPC 18:2 was assayed in plasma^30^. After ICB treatment in *CTLA-4^h/h^* mice, conversion of DLPC to LPC 18:2 was significantly diminished, consistent with reduced LPC 18:2 biosynthesis. (**Fig 3c**).

We next examined an acute model of dextran sulfate sodium (DSS)-induced colonic inflammation and colitis^31^ (**Fig 3a**, *bottom*). Unsaturated 18-carbon LPC are consistently depleted in the blood of DSS-treated mice, and are suggested to be functionally protective in colitis pathology^32^. Importantly, the precise identity of protective LPC and the mechanism of action are not well understood. Circulating LPC 18:2 concentrations significantly decreased with development of colitis mice after DSS treatment (**Fig 3d**), similar to as observed in ICB-treated *CTLA-4^h/h^* mice (**Fig. 3c**) as well as ICB-irAE patients (**Fig. 1**). Furthermore, LPC 18:2 biosynthesis was significantly diminished in the blood of DSS colitis mice (**Fig 3e**). In tumor-bearing mice, the pathology of DSS colitis is exacerbated by ICB treatment yet uncoupled from the anti-tumor response, as demonstrated by the systemic administration of TNF blockers that selectively neutralize ICB-irAEs without affecting tumor regression response^33^. In addition to TNF, the inflammatory pathology of DSS colitis also depends upon IL-6^34^, a pro-inflammatory cytokine that specifically promotes ICB-irAEs in patients independently of anti-tumor response^35^. We tested whether pro-inflammatory cytokines are involved in reduction of LPC 18:2 biosynthesis in inflamed mice. Neutralizing antibodies against TNF or IL-6 were administered systemically as a single dose, 7 days after DSS treatment and well after the onset of colitis pathology. Anti-IL-6 specifically restored LPC 18:2 biosynthesis in the blood, while anti-TNF had no effect (**Fig 3e**). Furthermore, in healthy individuals (**Supplementary Table 4**), IL-6 concentrations were found to be negatively correlated to LPC 18:2 (**Fig 3f**). Together, data show that inhibition of *de novo* biosynthesis of LPC 18:2 occurs in relation to inflammation, downstream of the pro-inflammatory cytokine IL-6.

### *In vivo* LPC 18:2 supplementation suppresses ICB-irAEs and colitis

To examine the biological function of LPC 18:2 *in vivo* and its causal relation to ICB-irAEs, purified LPC 18:2 was co-administered with ipilimumab and anti-PD-1 to *CTLA-4*^h/h^ mice, using intraperitoneal injection to bypass enteric uptake and metabolism (**Fig 4a, top**). In the supplemented mice, LPC 18:2 levels were restored in blood after intraperitoneal administration (**Extended Data Fig 5b**). Histology scoring was used to grade colon pathology in combination ICB-treated *CTLA-4^h/h^* mice (**Fig 4b, (Extended Data Fig 5c**). Supplementation with LPC 18:2 attenuated the development of colitis after combination ICB treatment (**Fig 4b, Extended Data Fig 5c**), suggesting that circulating LPC 18:2 limits the severity of ICB-driven irAEs *in vivo*. The effects of LPC 18:2 supplementation in reducing colon inflammation was also observed in DSS-treated colitis mice (**Fig 4c**), with diminished weight loss and colon shrinkage (**Extended Data Fig 6a-b**), a measure of fibrosis, after DSS treatment of LPC 18:2 supplemented mice. In contrast to the anti-inflammatory effect of LPC 18:2 *in vivo*, supplementation with LPC (16:0), structurally related molecules, including LPC (18:0) or LPC (18:1), or short-course anti-IL6 or anti-TNF treatment did not significantly affect colon pathology (**Extended Data Fig 6c-e**). In wild type mice bearing B16 melanoma or MC38 colon carcinoma tumors, circulating LPC 18:2 concentrations were maintained after combination anti-CTLA-4 and anti-PD-1 treatment in the absence of irAEs, and additional LPC 18:2 supplementation had no effect upon ICB-driven tumor regression (**Extended Data Fig 7a-b**). These data indicate that LPC 18:2 supplementation suppresses ICB-driven irAEs *in vivo*, without altering ICB-driven anti-tumor response.

**Figure 4:**
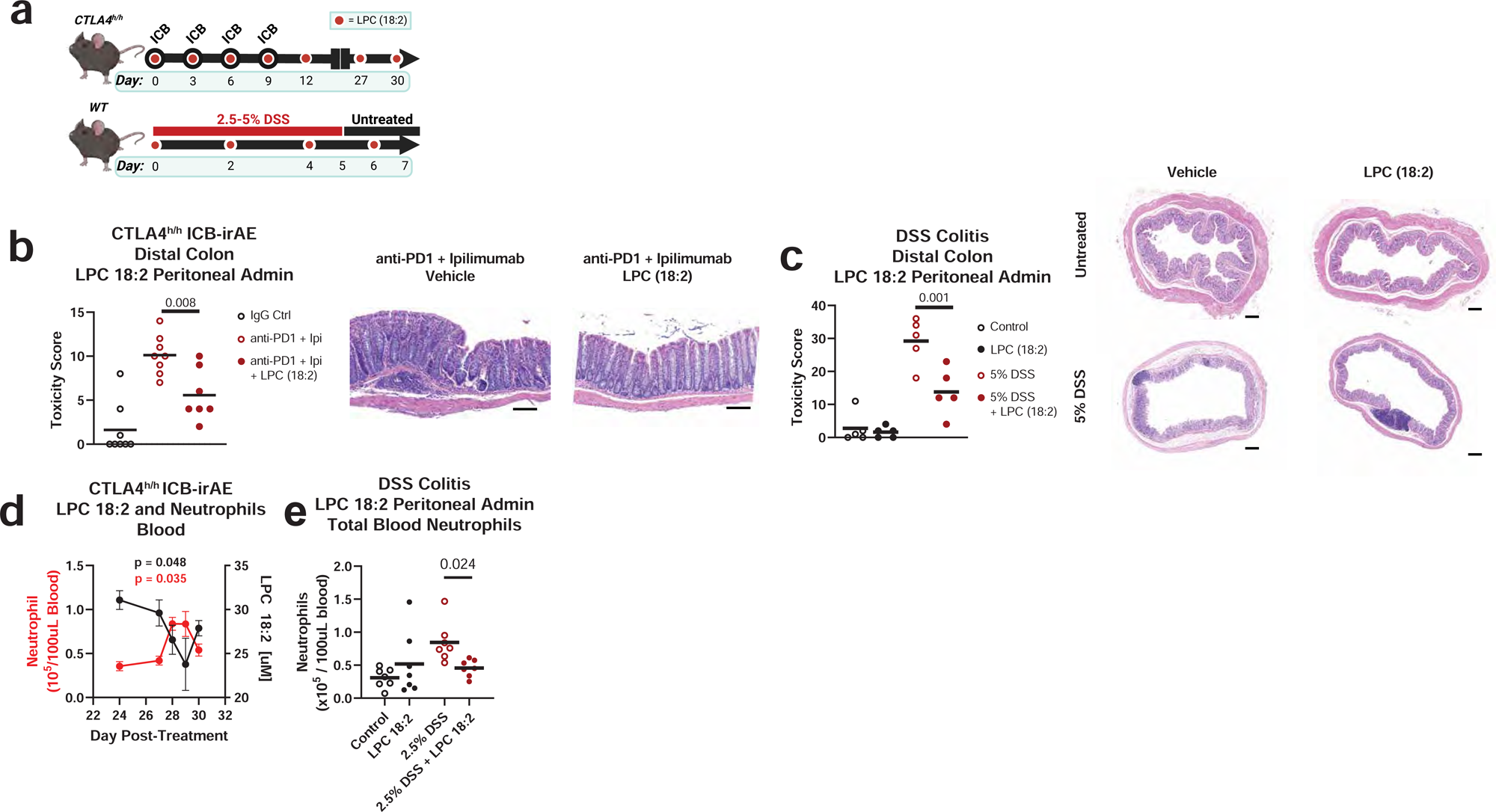
LPC 18:2 supplementation reduces colitis and neutrophilia. (**a**) Diagram of experimental design for LPC 18:2 intraperitoneal administration in ICB-treated *CTLA-4^h/h^*mice and DSS colitis. (**b**) Histology score and representative H&E staining in distal colon of *CTLA-4^h/h^* mice following intraperitoneal administration of 100 ug anti-human IgG1 isotype control (hIgFc) or 100 ug ipilimumab + 100 ug anti-mouse PD-1 monoclonal antibody (ICB), dosed as above, n = 7-8 per group. Error = SD, Bar = mean, ruler = 100µm (**c**) Histology score and representative H&E staining in distal colon of DSS-treated mice. LPC 18:2 was administered at 25 mg/kg on Days 0, 2, 4, and 6 of DSS treatment. n = 5 per group. Bar = mean, ruler = 200µm. (**d**) Mean LPC 18:2 concentration and neutrophil (CD45^+^, CD11b^+^, Ly6G^+^) numbers in *CTLA-4^h/h^* mouse blood. n = 5 per group. Error = SEM. (**e**) Multi-color flow cytometry of blood neutrophils in 2.5% DSS-colitis mice following treatment with 25 mg/kg of LPC 18:2. One-way ANOVA with Dunnett’s multiple comparisons test (Fig. 4b); two-way ANOVA with Sidak multiple comparisons test (Fig. 4c**, 4e**).

### LPC 18:2 correlates with reduced neutrophil counts and blunts colitis-associated neutrophilia

The correlation between lower circulating LPC 18:2 concentrations and higher peripheral blood neutrophil numbers observed in humans (**Fig 2**) suggests that LPC 18:2 may exert anti-inflammatory effects by regulating neutrophils in the blood. Similar to humans, *CTLA-4^h/h^* mice experience elevated blood neutrophilia after ICB treatment^46^ (**Extended Data Fig 5d**), though an effect on peripheral neutrophil count is not observed with chronic LPC 18:2 supplementation. Longitudinal analysis of serial blood from combination ICB-treated *CTLA-4^h/h^* mice confirmed that the onset of neutrophilia coincided with a reduction in LPC 18:2 concentrations (**Fig 4d** and **Extended Data Fig 8a**). To examine the causal relationship between LPC 18:2 and neutrophilia *in vivo*, we tested whether LPC 18:2 supplementation blunts neutrophilia. LPC 18:2 supplementation in DSS colitis mice, which experience profound neutrophilia, resulted in a significant decrease in peripheral neutrophil numbers (**Fig 4e**). Taken together, results show that circulating LPC 18:2 suppresses severe ICB-irAEs by maintenance of peripheral neutrophil homeostasis (**Fig 5**).

**Figure 5.**
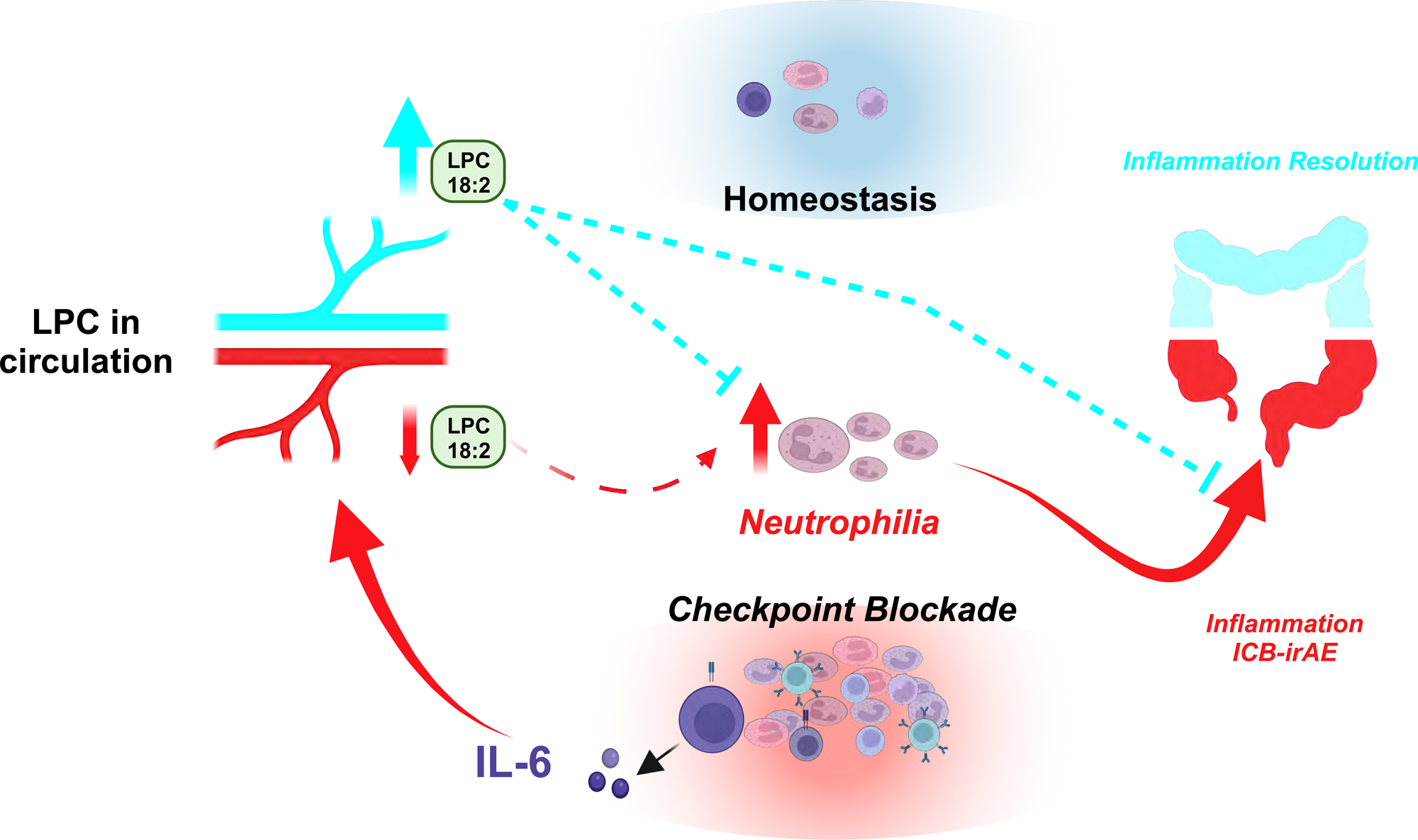
LPC 18:2 suppresses irAEs by regulating peripheral neutrophil homeostasis. Schematic representation of irAEs in ICB patients, which are linked to changes in peripheral LPC 18:2 biosynthesis.

## Discussion

Lower concentrations of circulating LPCs have been detected in prospective^36^ and prevalent^37^ blood sampling of myocardial infarction, diabetes, SLE, and sepsis. However, the exact LPC species involved and their functional role in regulating inflammation have remained unclear. Here we show that loss of circulating LPC 18:2 after ICB therapy precedes the development of severe grade III/IV ICB-irAEs across multiple organ systems. In ICB-irAE patients and healthy humans, lower LPC 18:2 concentrations in blood associated with higher neutrophilia. Loss of LPC 18:2 and dysregulated LPC 18:2 biosynthesis in the blood was also observed in ICB-irAE mice and colitis mice, and LPC 18:2 supplementation *in vivo* suppressed neutrophilia and irAE severity without affecting ICB anti-tumor activity. Together, these results point to a previously unrecognized role for LPC 18:2 in suppressing ICB-associated inflammation, and suggest that specifically targeting ICB-associated toxic inflammation as a therapeutic strategy could improve clinical ICB outcomes.

ICB-irAEs are increasingly appreciated as a broad descriptor of a group of immune-related toxicities, and are currently managed individually at the sub-specialty level^6^. In this study loss of circulating LPC 18:2 was broadly associated with several different types of ICB-irAEs as well as human autoimmune disease affecting multiple organ systems. This generalizability is consistent with the proposed function of LPC 18:2 in reducing pro-inflammatory neutrophils in the blood, which was a general feature of severe ICB-irAEs in our study and others^23^. However, blood neutrophilia alone is likely necessary but not sufficient to drive severe organ-specific damage, and future studies assessing LPC 18:2 and other bio-active molecules in affected organs will reveal the molecular determinants of ICB-irAE localization to specific organs. For example, infiltration of neutrophils from the blood into inflamed organs occurs due to tissue-specific pro-inflammatory signals produced by infection or damage. Pro-inflammatory signals drive local activation and permeabilization of the vascular endothelium, up-regulation of chemotactic ligands for the neutrophil CXCR2 receptor on inflamed cells, and enhanced survival and activity of migrating neutrophils. At the inflamed site, neutrophil effector functions depend on microenvironment-specific metabolic cues, local cytokine production and other tissue-derived signals^38^. For example, in cystic fibrosis lung infiltrating neutrophils acquire unique metabolic adaptations that sustain survivability via increased expression of glucose transporters that augment pro-survival pathways through CREB and mTOR and enhance lung inflammation^39^. Whether patients who develop pulmonary ICB-irAEs are enriched for similar metabolic adaptations is an intriguing possibility. Recent evidence indicates that organ-specificity of ICB-irAEs is linked to the type of cancer. For example, patients with melanoma, non-small-cell lung cancer or renal cell carcinoma are more likely to develop dermatological and gastrointestinal irAEs after treatment with anti-PD-1, as opposed to pneumonitis in patients with other solid tumor types^40^. Thus, it is likely that systemic inflammation in conjunction with tissue-specific pro-inflammatory signals contribute to the clinical diversity of organ-specific irAEs.

Despite the complexity and diversity of clinical ICB-irAEs, targeting the dynamic regulation of pro-inflammatory neutrophils in the blood through LPC 18:2 supplementation is likely to suppress multiple classes of severe ICB-irAEs. In the blood of ICB-irAE mice, the interval of LPC 18:2 decline coincided with a rise in phenotypically aged, CXCR2+ pro-inflammatory neutrophils and concomitant decrease in apoptotic cells. Studies of peripheral neutrophils exposed to purified LPC 18:2 further demonstrated cell-intrinsic effects in promoting superoxide-associated apoptosis, which is an important homeostatic mechanism for resolving inflammation *in vivo*^41^. In addition to direct effects on neutrophils, LPC 18:2 may regulate other immune cell populations in the blood to resolve neutrophil-driven inflammation, notably the phagocytic cells of the reticuloendothelial system, which promote clearance of aged, pro-inflammatory neutrophils in response to pro-resolving ROS signaling^41^, and eosinophils, whose abundance in peripheral blood has also been reported as predictive for ICB-irAEs^42^. Indeed, LPC are designated ligands for several different signaling receptors including G2A^43^, a G-protein coupled receptor associated with inflammation resolution in sepsis^44^ and colitis models^45^ that is expressed on neutrophils, eosinophils, myeloid cells and many other peripheral immune cell populations. Other LPC-derived lipids might also be involved in regulating ICB-irAEs. While our human studies specifically detected changes in LPC species, the enzymatic conversion of LPC to LPA, a family of bio-active lysolipids with well-characterized immune evasive signaling function^46^, is a consideration. Thus, in-depth studies of LPC receptor signaling on immune cells and exploration of other bio-active LPC-derived metabolites such as LPA are important considerations for further understanding the molecular regulation of ICB-irAEs.

Regarding the particular involvement of ICB-activated T cells, multiple lines of evidence point to a secondary or supporting role for adaptive immunity in many organ-specific irAEs. Although ICB-myocarditis and ICB-T1D resemble classical autoimmune disease, the more common ICB-irAEs including colitis, pneumonitis, hypophysitis and dermatitis are classified as autoinflammatory conditions, as they lack discernible involvement of self-antigens and auto-reactive T cells. To account for this diversity, we studied preclinical models for autoreactive T-cell dependent inflammation (e.g. ipilimumab-treated *CTLA4h/h* mice) and activated T-cell enhanced inflammation (e.g. DSS colitis). In both models, loss of LPC 18:2 in blood was observed downstream of inflammation and LPC 18:2 supplementation reduced the severity of toxic inflammation *in vivo*. The precise role of ICB-activated T cells in this process remains unclear. It is possible that ICB-activated T cells in circulation simply produce or activate production of pro-inflammatory cytokines. In colitis mice, LPC 18:2 production was partially restored by a single dose of neutralizing anti-IL-6 antibody, administered well after the onset of inflammatory pathology. These data support a role for IL-6 in ICB-irAE pathology, and are in accordance with the established function of IL-6 in protecting peripheral neutrophils from apoptosis during inflammation *in vivo*^47^. Interestingly, sPLA2 induction has been observed in cancer patients treated with recombinant IL-6^48^. Interferon-γ and TNF have also been shown to modulate the Lands Cycle via regulation of coenzyme-A independent transacylase (CoAIT) and LPCAT expression in macrophages *in vitro*^49^, supporting LPC change mediated by inflammatory cytokines. Indeed, coincidence of serum IL-6 and sPLA2 elevation in severe acute COVID-19^50^ supports a coregulatory mechanism in blood and potentially in liver, where acute phase reactants are often derived and PC acylases are dynamically regulated during inflammation. Thus, strategies to restore LPC 18:2 via supplementation or anti-IL-6 therapy might be explored after an irAE diagnosis to control inflammatory neutrophils in the blood and suppress deleterious inflammation.

## Online Methods

### Materials & Cell Lines

Purified LPC (16:0), LPC 18:0, LPC 18:1, and LPC 18:2 were purchased from Avanti Polar Lipids. Dextran sulfate sodium was purchased from Affymetrix. Monoclonal anti-mouse CTLA-4 (CD152) and anti-PD-1 (CD279) antibody, as well as IgG2a isotype control antibody, were purchased from InVivoMAb. Clinical-grade ipilumumab was a kind donation from S.P.P. B16-F10 cells were a kind donation from Dr. Stephen Schoenberger (La Jolla Institute for Immunology) and MC38 mouse colon adenocarcinoma cell line provided by J Schlom (National Cancer Institute). Cell lines were maintained as previously described^51^.

### Cohort Information

Archived and de-identified blood plasma or whole blood samples were obtained in accordance with the institutional review boards of each institute (University of Southampton, Yale University, University of California San Diego, Finnish Institute for Health and Welfare, HFGP and La Jolla Institute for Immunology IRB). Clinical practice from the American Gastroenterological Association and the National Comprehensive Cancer Network guidelines do not recommend special diets in management of the condition. Accordingly, no patients in the current study cohorts were instructed to do so.

### Southampton Ipilimumab

Cohort 1 is a retrospective study of ipilimumab monotherapy-treated metastatic melanoma patients with the Southampton University Hospitals NHS Foundation Trust (North West-Greater Manchester Central Research Ethics Committee, MREC number 12/NW/0157, NIHR portfolio adoption ID 18329). Patients of all sexes and ages greater than 18 years were identified via evaluation of a centralized prescribing system, and written informed consent was obtained prior to enrollment. Venous blood was collected directly before first intravenous administration of 3 milligrams per kilogram body weight ipilimumab, and then prior to subsequent ipilimumab administrations every three weeks. Blood plasma samples were matched against complete blood count with differentiation where available from the same venous sample. Covariates include patient age, sex, and sample matrix batch.

### Moores Cancer Center Mixed ICB

Cohort 2 is a community-acquired sample set acquired from solid tumor patients receiving PD-1, PD-L1, CTLA-4, or PD-1 and CTLA-4 combination therapies, derived from the University of California, San Diego Moores Cancer Center Biorepository (HRPP# 150348). All biorepository participants gave informed consent for their samples to be stored, and patient information was appropriately de-identified. Patients of all sexes and of age 18 years or older were included. Only patients for whom both baseline (fewer than one week prior to initiation of immune checkpoint blockade) and longitudinal blood plasma collections were available. Hematologic malignancies were excluded, as well as those where primary therapy included monoclonal antibodies against targets other than PD-1, PD-L1, or CTLA-4. Patient information was independently de-identified prior to storage. Covariates included age, sex, and sample matrix batch. Development of adverse events were recorded by study clinicians, with laboratory or hematologic abnormalities, events unlikely to be associated with intervention, or events unlikely to be immune mediated were excluded. Additionally, patient blood plasma was matched to complete blood count with differentiation data drawn, where such data were available from the same phlebotomy sample, for additional analysis.

### Yale Pembrolizumab

Cohort 3 is a two-part, non-randomized, parallel assignment interventional trial (NCT02407171) of pembrolizumab with single-target stereotactic beam radiotherapy (SBRT) in melanoma or non-small cell lung carcinoma, conducted by Yale University. Patients of all sexes and of age 18 years or older were eligible for enrollment; exclusion criteria included known immunocompromised or autoimmune conditions, monoclonal antibody therapeutics fewer than four weeks prior to enrollment, chemotherapy or radiation therapy fewer than two weeks prior, or tyrosine kinase inhibitors 72 hours prior. Patients with active or untreated CNS/leptomeningeal metastases were also excluded.

Enrolled participants were sorted into previous PD-1 treatment progression or treatment naïve. Treatment naïve patients were given 200mg IV pembrolizumab every two weeks until progression on therapy, at which time they were reallocated to receive concomitant SBRT treatment. Previous PD-1 treatment progressing patients were allocated to receive SBRT prior to initiating pembrolizumab. SBRT was given to a single site at total 3000 cGy dose in either 3 or 5 fractions; two study arms included lung intraparenchymal and extraparenchymal masses. Patient blood plasma was isolated directly prior to or at intervals following the initiation of pembrolizumab. Covariates included age, sex, and sample matrix batch. Development of adverse events were recorded by study clinicians, with laboratory or hematologic abnormalities, events unlikely to be associated with intervention, or events related to drug and/or SBRT but not immune mediated were excluded.

### 500FG

The 500FG cohort is a community-acquired healthy human subset of the Human Functional Genomics Project (Ethical Committee of Radboud University Nijmegen, the Netherlands, #42561.091.12). Participants of all sexes, over the age of 18 years were included. Venous blood samples were collected after informed consent was obtained. Flow cytometry, immunoglobulin quantification, and cytokine measurements were performed as described in previous studies^40,41^. Cells were analyzed on a Navios flow cytometer with solid-state lasers at 405, 488, and 638 nanometer wavelengths. Elucidation of 73 distinct cell populations was performed using Kaluza Software version 1.3, with hierarchical gating strategy outlined previously^41^. Immunoglobulin concentrations were determined by Beckman Coulter Immage against reference (ERM®-DA470). IL6 quantification was performed using a PeliKine Compact ELISA system (Cat M9316). Covariates included participant sex and age, as well as sample matrix batch.

### FINRISK-2002

The FINRISK-2002 cohort is a longitudinal sub-cohort of the Finland National FINRISK study (Coordinating Ethical Committee of the Helsinki and Uusimaa Hospital District, Ref. 558/E3/2001). Dietary associations are derived from a 48 hour recall survey subset (FinDiet 2002, n = 2007)^52^. Prevalent conditions from FINRISK-2002 utilized in this study include chronic inflammatory conditions (systemic lupus erythematosus, inflammatory bowel disease, asthma, and rheumatoid arthritis), and participants without a diagnosed medical condition at the time of last study record. Individuals of all sexes and age of greater than 24 years were included, and informed consent was obtained prior to venous blood collection. Blood plasma samples were matched to complete blood count with differential, immunoglobulin, or cytokine data where data from the same phlebotomy sample where available^73^. Covariates include patient sex, age, and sample matrix batch.

### Bioactive Lipid Extraction from Blood Plasma

Na-EDTA or heparin prepared blood samples were thawed from storage at -80°C overnight in light-free conditions at 4°C. All extractions were performed in 96-well format. 20uL of each sample were mixed with 80uL of -20°C ethanol containing 20 deuterated standards to precipitate protein and extract lipid content. Samples were vortexed at 4°C and 500rpm for 15 minutes and centrifuged for 10 minutes to sediment protein content. From each supernatant, 65uL were taken and mixed with 350uL water in an Axygen 500uL retention v-bottom 96 well plate. To improve extraction, an additional 65uL of -20°C ethanol was added gently to the protein pellet, vortexed gently by hand for 15 seconds, and added to the same well.

Complete extracted sample volumes were loaded onto a Phenomenex Strata-X 10mg/mL polymeric solid phase extraction (SPE) 96-well plate pre-washed stepwise with 600uL methanol, 600uL ethanol, and equilibrated with 900uL water. Following gravity elution from each SPE well, 600uL of 9:1 water:methanol were added as a wash, pulled through slowly at 5mmHg, and then increased to 20mmHg for 45 seconds to fully dry each SPE. Bound metabolites enriched for bioactive lipids were then eluted in 450uL ethanol into a fresh 500ul Axygen v-bottom plate. Sample eluate was dried in a vacuum concentrator at 40°C until completely dried, before adding 50uL per well of 75:20:5 water:methanol:acetonitrile containing 10uM CUDA as resuspension solvent. Sample plates were vortexed at 500 RPM for 10 minutes at 4°C to fully resuspend metabolites, before transferring to 300uL glass inserts in a 96-well Greiner deep well plate and immediately sealed. Samples were then immediately analyzed via LC-MS/MS.

### Liquid Chromatography-Tandem Mass Spectrometry

Directed, nontargeted liquid chromatography tandem mass spectrometry for the detection of bioactive lipid metabolites was performed as described previously^29^. LC-MS/MS was performed on a Thermo Vanquish UPLC system coupled to a Thermo QExactive Orbitrap mass spectrometer. Injection volume for each sample was 20uL onto a Phenomenex Kinetex C18 (1.7um particle size, 100 x 2.1 mm) column. Mobile phases were composed of; A: 70% water, 30% acetonitrile, 0.1% acetic acid, and B: 50% acetonitrile, 50% isopropanol, 0.02% acetic acid. Flow rate was a constant 0.375 mL/min, with gradient mobile phase as follows: 1% B from - 1.00 minutes to 0.25 minutes, 1% to 55% B from 0.25 minutes to 5.00 minutes, 55% B to 99% B from 5.00 minutes to 5.50 minutes, and 99% B from 5.50 minutes to 7.00 minutes. Column temperature was 50°C, with a 50:25:25:0.1 water:acetonitrile:isopropanol:acetic acid needle was set to 5 seconds post-draw. Mass detection was performed with an equipped heated electrospray ionization (HESI) source with manually optimized source geometry^29^. Negative mode profile data was acquired for all samples, with sheath gas flow, aux gas flow, and sweep gas flow of 40, 15, and 2 units, respectively. Spray voltage was -3.5kV, and capillary and aux gas temperature were 265 and 350°C, respectively, with S-lens RF at 45. MS1 scan events were in a scan range of *m/z* 225-650, mass resolution of 17.5k, AGC of 1e6 and inject time of 50ms. To assist quantification and aligning intra- and inter-cohort chromatographic drift, tandem mass spectra were acquired using collision-induced dissociation (CID). Data independent acquisition (DIA) acquired in the following four mass windows: m/z 240.7–320.7, m/z 320.7–400.7, m/z 400.7–480.7, and m/z 480.7–560.7, with a mass resolution of 17.5, and AGC of 1e6, and an inject time of 40ms. Metabolite matching to LC-MS features was performed against an in-house library of bioactive lipids or by matching high quality tandem mass spectra against target features when identified.

### LC-MS Data Handling

LC-MS peak identification was performed using deep neutral network-based classification as described previously^30^. Briefly, Thermo raw to mzXML file conversion was performed using MSconvert version 3.0.9393 (ProteoWizard), from which initial bulk LC-MS feature alignment was performed using in-house, R-based landmark identification and retention time correction. Chromatographic drift-corrected mzXML were then converted into composite raster image files including m/z and retention time windows bounding putative features. High-confidence features were then identified using a trained neutral network specific for this LC-MS/MS method. Semiquantitative comparisons of LC-MS features utilized peak height intensity values.

### Animal Handling

Wild type C57BL/6 mice were purchased from Jackson Laboratories and *CTLA-4^h/h^* mice^46^ obtained from the Institute for Human Virology at University of Maryland or purchased from Genoway. Mouse experiments were performed with animals housed in pathogen-free conditions at the La Jolla Institute for Immunology or at the Institute for Human Virology at University of Maryland. All procedures involving mice were performed according to the respective Institutional Animal Care and Use Committee at LJI or University of Maryland.

### Dextran Sulfate Sodium-Induced Colitis Model

Colitis was induced in 9-12 weeks C57BL/6 mice (purchased from Jackson Laboratories) using one cycle of either 2.5% or 5% dextran sulfate sodium (DSS) (Affymetrix, Chem-Impex) treatment in drinking water. Same sex littermates were utilized; data from male mice, in which colitis was more robustly induced, are reported here, with results validated in female mice. Mice were given 2.5 or 5% DSS as indicated for 5 days, followed by two days on untreated water. Primary humane experimental endpoint was body weight loss in excess of 20% starting weight, measured daily. In some cases, mice were administered intraperitoneally with either control saline solution or indicated concentration of LPC. Severity of colitis was measured at experiment termination by body weight loss, colon length, and histological criteria (below). Where indicated, blood was collected in EDTA-coated Eppendorf tubes from terminal mice by cardiac puncture, with cervical dislocation as secondary euthanasia. Blood was immediately centrifuged at 2000 RPM and 4°C for 20 minutes to collect blood plasma, which was stored at -80°C until analysis.

### Human CTLA-4 Mice

Mice expressing chimeric CTLA-4 (*CTLA-4^h/h^*), containing the extracellular domain of human CTLA-4 within the context of endogenous mouse *Ctla4* locus backcrossed onto the C57BL/6 background, have been described previously^46^ and were obtained the Institute for Human Virology at University of Maryland or from Genoway. Multi-organ irAEs after ipilimumab treatment of these mice, affecting the heart, liver, lung, kidney, salivary gland and colon, have been extensively characterized. Same sex littermate mice were assessed, with data from female mice presented due to increased irAE severity compared to male mice. ICB antibodies and LPC 18:2 were administered intraperitoneally at the indicated dose and time interval. Primary humane experimental endpoint was body weight loss in excess of 20%, measured every third day. Mice were humanely euthanized for toxicity scoring and blood plasma LC-MS analysis on Day 30. Complete blood counts were collected on Day 29 (penultimate) of treatment from 50 uL of blood, collected in EDTA-coated Eppendorf tubes and analyzed by HEMAVET HV950 blood analyzer (Drew Scientific), according to manufacturer’s protocol, or by flow cytometry.

### Histology

Multi-organ ICB toxicity scores in *CTLA-4^h/h^* mice were generated from histology scoring of the heart, lung, salivary gland, colon, and liver as described previously by Du *et al*^25^. Distal colon or cecum toxicity was determined from hematoxylin and eosin stained tissues as described. Briefly, cecum and distal colon samples were collected and fixed in zinc formalin (Medical Chemical Corporation) for at least 24 hours prior to paraffin embedding. Tissue was stained with hematoxylin and eosin, and at least six representative 5um slices were collected from each tissue for embedding on slides. Image acquisition was performed on an Axioscan Z1 platform (Zeiss), using a 20x objective lens, utilizing the Zen 2.3 software automatic scan mode. Slides were blinded and then scored on a composite of the following criteria: inflammation (0 = none, 1 = mild, 2 = moderate, 3 = severe); infiltration (0 = none, 1 = mucosal or submucosal, 2 = mucosal and submucosal, 3 = transmural); crypt damage (0 = none, 1 = basal 1/3 damaged, 2 = basal 2/3 damaged, 3 = only surface epithelium intact, 4 = entire crypt and epithelium lost); and edema (0 = none, 1 = 1.5-2x submucosal thickness, 2 = >2x submucosal thickness)^53,54^. Four randomly selected areas were analyzed and a combined histological score was determined by adding the scores of the individual sections.

### Quantification of LPC Production from Diacylphosphocholine

LPC production was determined via an adaptation of the method delineated by Mouchlis *et al*^30^. Where indicated, rat anti-mouse IgG1 monoclonal antibodies against TNF alpha (MP6-XT22), IL-6 (MP5-20F3), or a control antigen (eBRG1) (eBioscience) were administered intraperitoneally six hours prior to isolation for acute cytokine inhibition. Liver or fresh blood plasma were harvested and fresh frozen at -80°C prior to analysis. On the day of analysis, liver lysate was prepared using bead beater homogenization with cold 50mM Tris-HCl, pH 7.5 until homogenous, followed by 5000 xg centrifugation at 4°C for 15 min to pellet insoluble material. Working samples were then prepared at 0.5ug/mL or 1ug/mL protein on ice for liver lysate and blood plasma, respectively. Test article (18:2 (Cis) PC (1,2-dilinoleoyl-sn-glycero-3-phosphocholine or DLPC), Avanti Polar Lipids) was prepared in 4mM C_12_E_8_ and diluted to 1mM DLPC in each sample preparation. Sample with test article were prepared in technical triplicate for 30min and 1hr incubations at 37°C, before reactions were quenched with ice-cold LC-MS grade ethanol before analysis by tandem LC-MS as above. LPC 18:2 *de novo* production is represented as nanomoles/L/min LPC 18:2 in an average of technical triplicates after subtracting rates from both sample and test article controls.

### Flow Cytometry

Please see Supplementary Materials for all fluorochrome-conjugated antibodies and dyes with staining conditions used here. For flow cytometry parameters used in analysis of the 500FG cohort, please see Aguierre-Gamboa *et al*^22^. Mouse peripheral blood cellularity were determined from retro-orbitally acquired venous blood with red blood cell lysis prior to staining.

To assess the immune cell populations and their apoptotic state, the following flowcytometry protocol was followed. RBC lysis was performed on isolated venous blood using RBC lysis buffer (Biolegend), after which live cells were stained with the LIVE/DEAD Fixable Blue Dead Cell Stain (Invitrogen) for 30 min on ice and washed twice with FACs Buffer (1xPBS containing 2% FBS and 0.05% sodium azide). Next, Fc receptors were blocked with unconjugated anti-CD16/32 (BD Pharmingen) for 15 min on ice prior to staining with the following antibodies: APC-Cy7 labeled CD45 (Clone: 30-F11, Biolegend), BV605 labeled CD11b (Clone: M1/70, Biolegend), PerCP-Cy5.5 labeled Sig F (Clone: S17007L, Biolegend), BV711 labeled CXCR4 (Clone: L276F12, Biolegend), BV570 labeled Ly6C (Clone: HK1.4, Biolegend) and PE/Dazzle labeled Ly6G (Clone: 1A8, Biolegend) for 30 min on ice in the dark. Subsequently, samples were washed twice with FACs buffer and fixed using 4% PFA. In order to stain for apoptotic neutrophils, post-extracellular staining, samples were washed twice with the Annexin V binding buffer (Biolegend) and stained using the Pacific Blue labeled Annexin V (Biolegend) as per instructions provided by the manufacturer (15 minutes at room temperature in the dark). Acquisition was performed using the BD Fortessa flow cytometry systems with associated acquisition software. Population gating was done using FlowJo software.

### In vitro neutrophil apoptosis and ROS production

Peripheral human neutrophils were isolated from whole blood using the Miltenyi Biotec’s negative isolation kit - MACSxpress-Whole Blood Neutrophil Isolation kit as per the manufacturer’s instructions. RBC lysis buffer (Biolegend) was used to eliminate contaminating RBCs. Isolated neutrophils were then stored at 37°C in RPMI media (Gibco) containing 5% FBS. To validate the effects of LPC presence or absence on human neutrophil activation and apoptosis, the following assay was performed. Briefly, isolated neutrophils were washed with RPMI media (serum free media) and then incubated for 10 min with 10µM dihydrorhodamine 123 (DHR) (Invitrogen). Samples were then treated with LPC 18:0, LPC 18:2, Leukocyte activation cocktail (BD Pharmingen) or the vector control (media) and collected for flowcytometric analysis at 30 and 120 minutes post-treatment. Collected samples were then stained as follows. Live cells were stained with the LIVE/DEAD Fixable Blue Dead Cell Stain (Invitrogen) for 30 minutes on ice and washed twice with FACs Buffer. Samples were then washed twice with the Annexin V binding buffer (Biolegend) and stained using the Pacific Blue labeled Annexin V antibody (Biolegend) as per instructions provided by the manufacturer (15 min at room temperature in the dark). Acquisition performed using the BD Fortessa flow cytometry systems with associated acquisition software. Population gating was done using FlowJo software.

### Subcutaneous Tumor Modeling

C57BL/6 mice were used to model tumor growth and anti-tumor ICB efficacy. B16-F10 melanoma cells (1E6) were seeded in subcutaneous flanks in 9-12 weeks old mixed sex littermate mice. Tumor volume was measured as (A x B^2^)/2, where A = the largest and B = the smallest diameter by caliper. Primary humane experimental endpoint was body weight loss in excess of 20% or tumor volume in excess of 2000mm^3^. Where indicated, 100 ug anti-mouse CTLA-4 and 100 ug anti-mouse PD-1 or 200 ug IgG2 isotype control antibody (InVivoMAb), or 25 mg/kg LPC 18:2, were administered intraperitoneally in 100 µL saline on Days 4, 7, 11, and 14 following tumor cell injection.

### Statistical Analysis

Associations between changes in bioactive lipid abundance and irAE severity were handled as ordinal logistic regressions between “none”, “Grade I/II”, and “Grade III/IV” groups; anti-tumor response associations were similarly handled as ordinal according to RECIST 1.1 metrics^35^ of efficacy. Maximum excursion was treated as the largest absolute magnitude fold change of an LC-MS feature peak height relative to patient’s baseline sample following base-2 logarithmic transformation. Regressions included patient age and sex as covariates.

For continuous variables, including PBMC cellularity, linear regressions were performed against base-10 log transformed variables, with age, sex, and body mass index (BMI) as covariates.

For in vitro analysis of neutrophils, data values are reported as mean +/- standard deviation, with differences determined using two-tailed Student’s t-test. Regarding statistical calculations, normal data distribution was confirmed using controls or previously generated large-scale datasets (n>384) prior to analysis of experimental samples. For *in vivo* and *in vitro* experiments, all reported results were independently replicated using three independent experiments.

## Data Availability

All data supporting the findings of this study, including patient information and demographics, LC-MS feature quantitative measurement and raw LC-MS data, are contained within the article or available within Supplementary Information.

## Acknowledgements

We thank Dr. Vijayanand Pandurangan for input on the Southampton cancer patient cohort; Mehak Kaur for assistance with *in vivo* mouse models; Dr. Zbigniew Mikulski, Katarzyna Dobaczewska and Angela Denn and the LJI Histology and Microscopy Core for expertise in tissue handling and processing; Dr. Daniel Giles for support with DSS colitis model; Dr. Roy Decker for access to the Yale cohort; Mahan Najhawan, Lily Quach, Thien-Tu Catherine Nguyen for mass spectrometry expertise; Maija Corey for work on primary human neutrophils. This work was supported by National Institutes of Health (NIH) grant nos U01DE028227 (J.S.G., S.S.), U54AG065141 (M.J., S.C., S.S.) and R01CA273230 (S.K., S.C., M.J., S.S.); S10OD020025 and R01ES027595 (M.J.); NIH P01DK46763 (M.K.); NIH U01CA233364 (A.A.P.); T32GM007752 and F31CA236405; a Tulie and Rickie Families SPARK award (I.T.M.) and an American Cancer Society Postdoctoral Fellowship PF-20-132-01-LIB (M.A.M.). M.G.N. was supported by an ERC Advanced grant (833247) and Spinoza grant of the Netherlands Organization for Scientific Research. T.N. was supported by the Research Council of Finland (Grants 321351 and 354447) and the Sigrid Jusélius Foundation.

## Author Information

Contributions: I.T.M., M.J. and S.S. conceived and designed the study. P.J., A.S.H., T.N., V.S., S.J.C., A.M.C., S.P.P., S.M.K., C.O., A.A.P., L.A.B.J, M.G.N. and P.V. provided or collated patient sampling and metadata, I.T.M. and K.D. acquired and assembled the mass spectrometry metabolomics data. I.T.M., M.H., J.D.W., T.L. and S.C. analyzed, integrated and interpreted the mass spectrometry metabolomics and orthogonal immune signature data. P.S. contributed to study design, optimized and conducted irAE mouse model studies, performed and analyzed immune phenotyping of peripheral blood cells, and optimized and implemented sample collection and analysis of human neutrophils. C.F. performed sample collection and flow cytometry analysis of human neutrophils. N.N. performed and analyzed immune phenotyping (flow cytometry analysis) of peripheral blood cells from irAE models. M.H. and P.Z. performed and analyzed LPC supplementation in *CTLA-4*^h/h^ mice. S.J.G. provided expertise for GPCR signaling. M.D. and M.K. provided support for executing and analyzing DSS colitis mice. M.M. and C.C.H. provided expertise for neutrophil gating, phenotyping and functional studies. S.S. and M.J. provided overall direction and supervision for the project. I.T.M, M.J. and S.S. wrote the manuscript with input from co-authors.

## Ethics Declarations

We have complied with all relevant ethical regulations. For the human studies, all subjects gave their informed consent for inclusion before they participated in the study. S.T., J.D.W, M.J. and S.S. have financial interest in Sapient Bioanalytics, LLC.

## Code Availability

Liquid chromatography tandem mass spectrometry data were collected using Thermo Scientific proprietary software for Vanquish UHLPC and QExactive MS systems. Logistic and linear regression analyses of LC-MS data were performed using R coding software.

## Extended Data Legends

**Extended Data 1.**
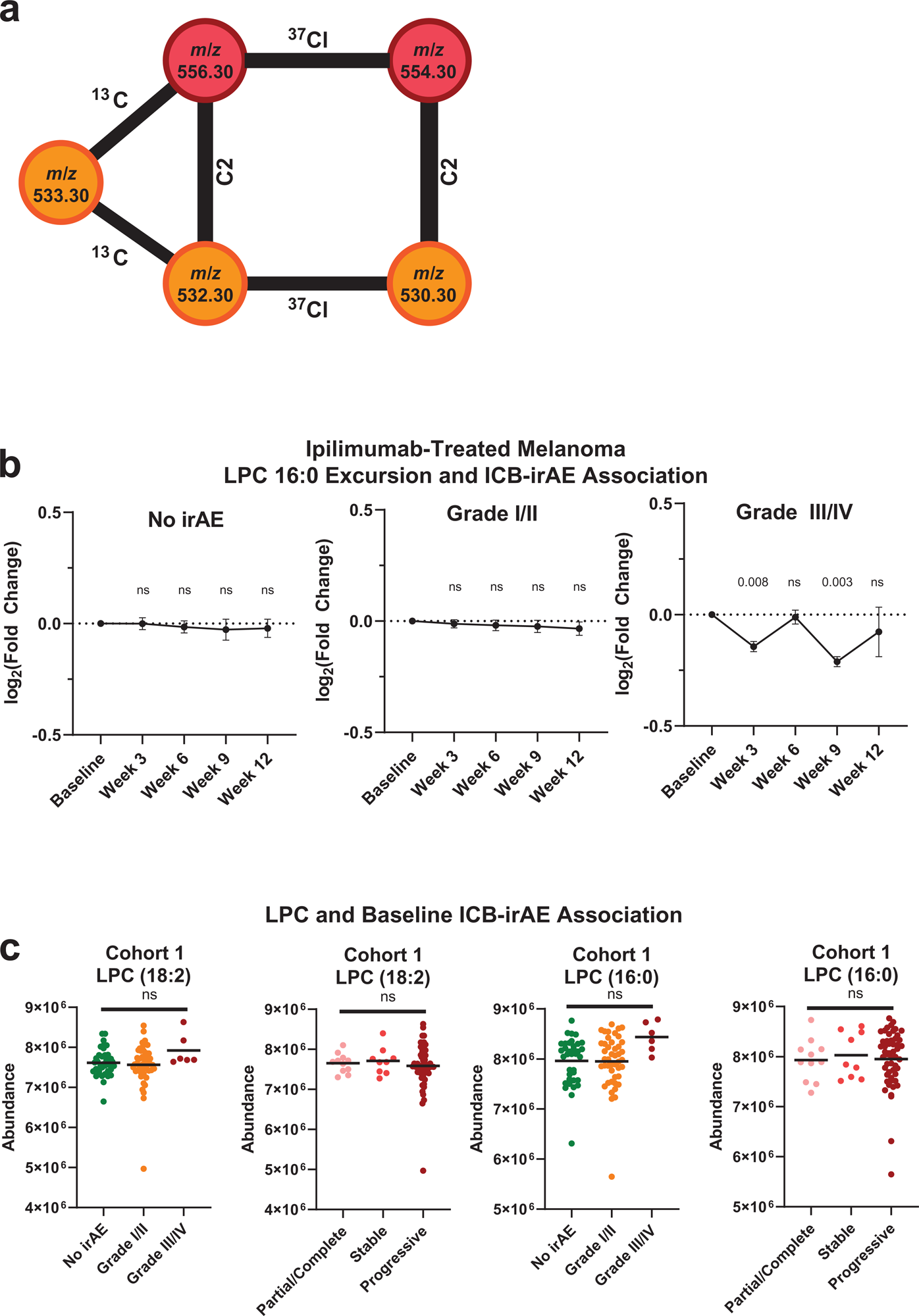
LPC 18:2 and LPC 16:0 association with ipilimumab-treated melanoma patient outcomes. (**a**) Tandem mass spectral chemical network of 5 irAE-associated LC-MS features corresponding to [M+Cl^-^] adducted LPC 18:2 and LPC 16:0. (**b**) Excursion of LPC (16:0) in cohort 1 ipilimumab treated patient blood plasma (n = 65). Error = SEM. (**c**) Concentration of LPC 18:2 and LPC (16:0) in baseline sampling from cohort 1.

**Extended Data 2.**
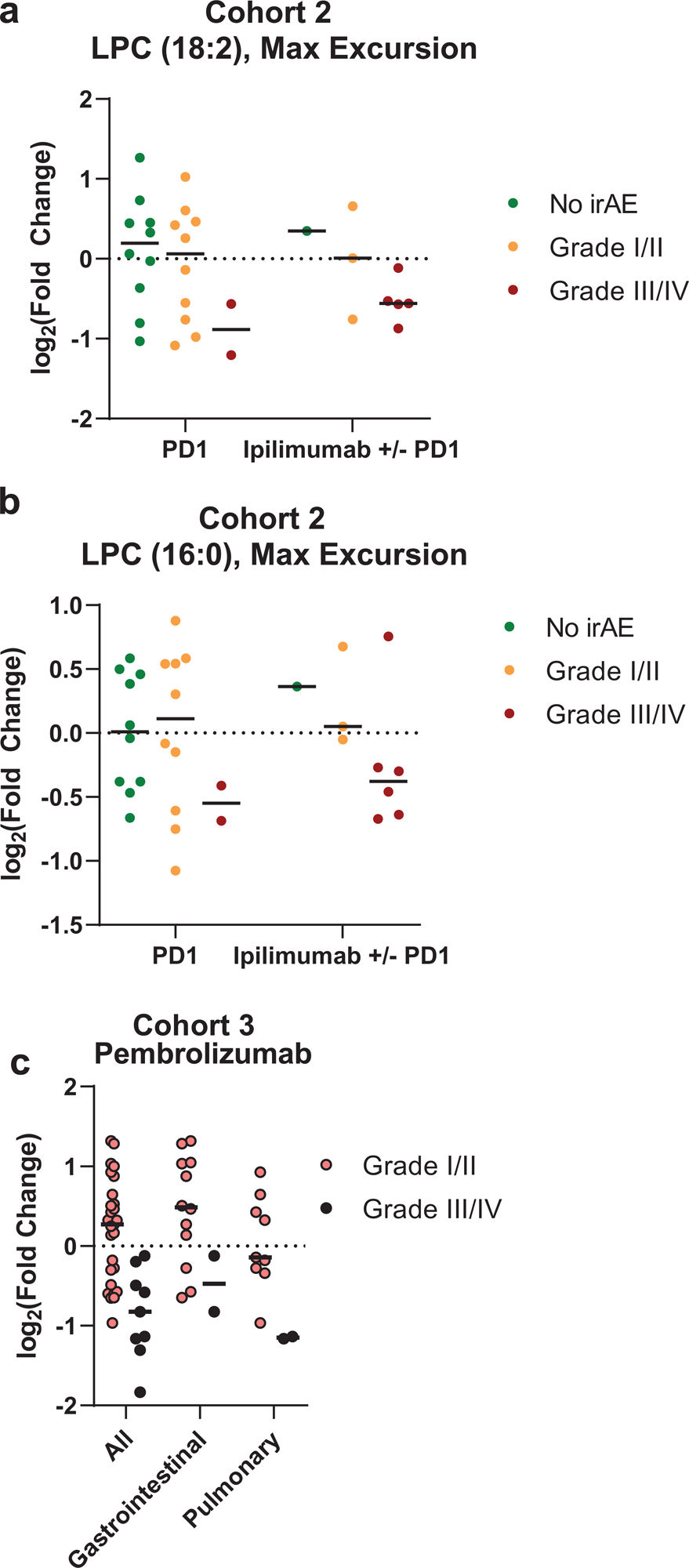
LPC 18:2 and LPC 16:0 excursion in different ICB regimens. (**a-b**) Maximum excursion of LPC 18:2 (**a**) and LPC 16:0 (**b**) in cohort 2, broken out by anti-PD-1 monotherapy or anti-PD-1 combination ipilimumab (n=65). (**c**) Maximum excursion of LPC 18:2 in cohort 3 among patients with ICB-irAE, bars = mean. One-sided Student’s t-test (**Extended Data 2c**).

**Extended Data 3.**
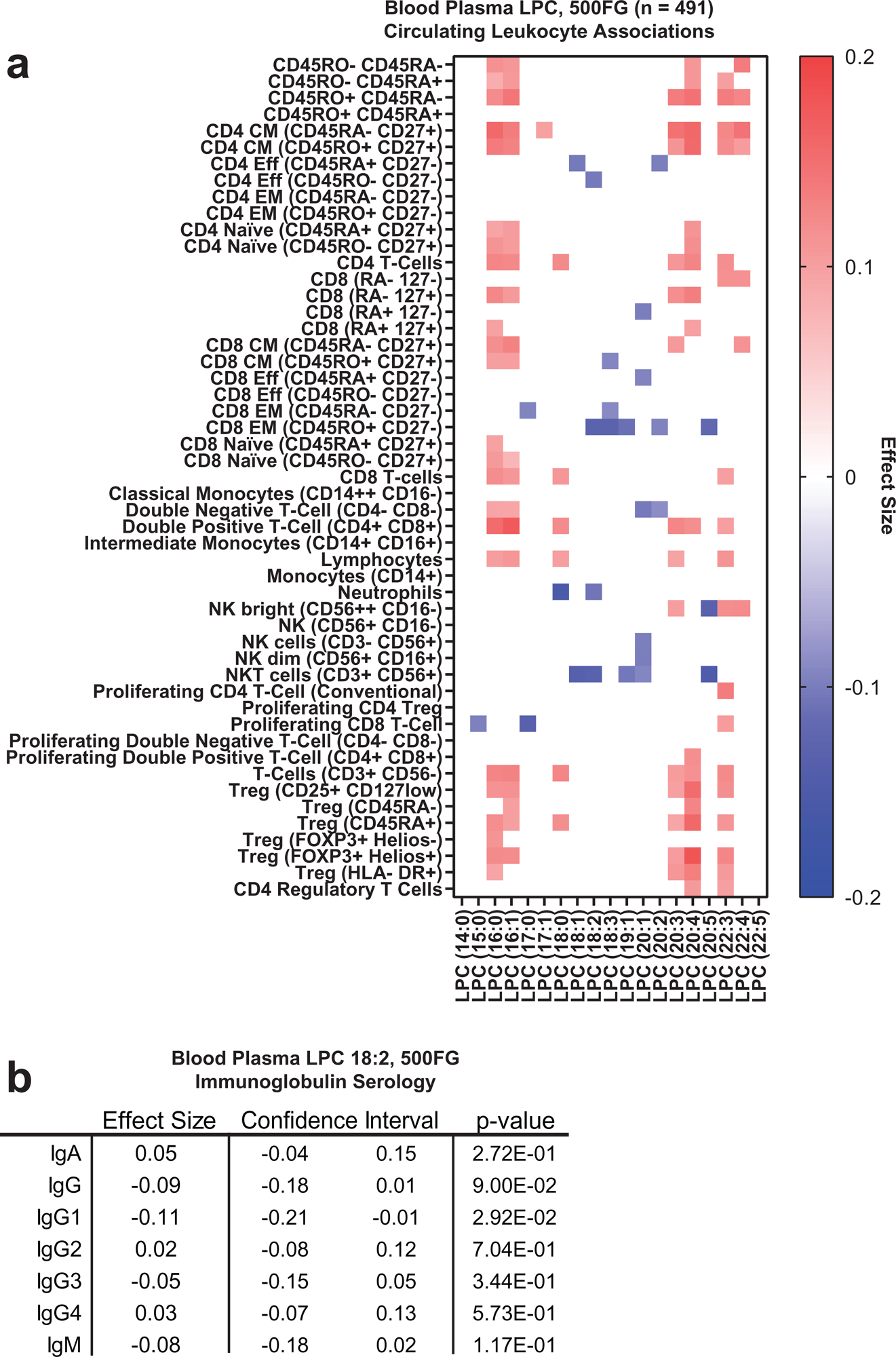
LPC associations with immune signatures in 500FG and FINRISK 2002 community-dwelling cohorts. (**a**) Heat map of LPC associations with immune cellularity by multi-color flow cytometry in 500FG (n = 488). Color = effect size with p < 0.05. (**b**) Circulating LPC 18:2 associations by linear regression with serum immunoglobulin titers in 500FG (IgG [7.00-16.00 gram/Liter]; IgA [0.70-4.00 gram/Liter]; IgM [0.40-2.30 gram/Liter]; IgG1 [4.90-11.40 gram/Liter]; IgG2 [1.50-6.40 gram/Liter]; IgG3 [0.20-1.10 gram/Liter]; IgG4 [0.08-1.40 gram/Liter].

**Extended Data 4.**
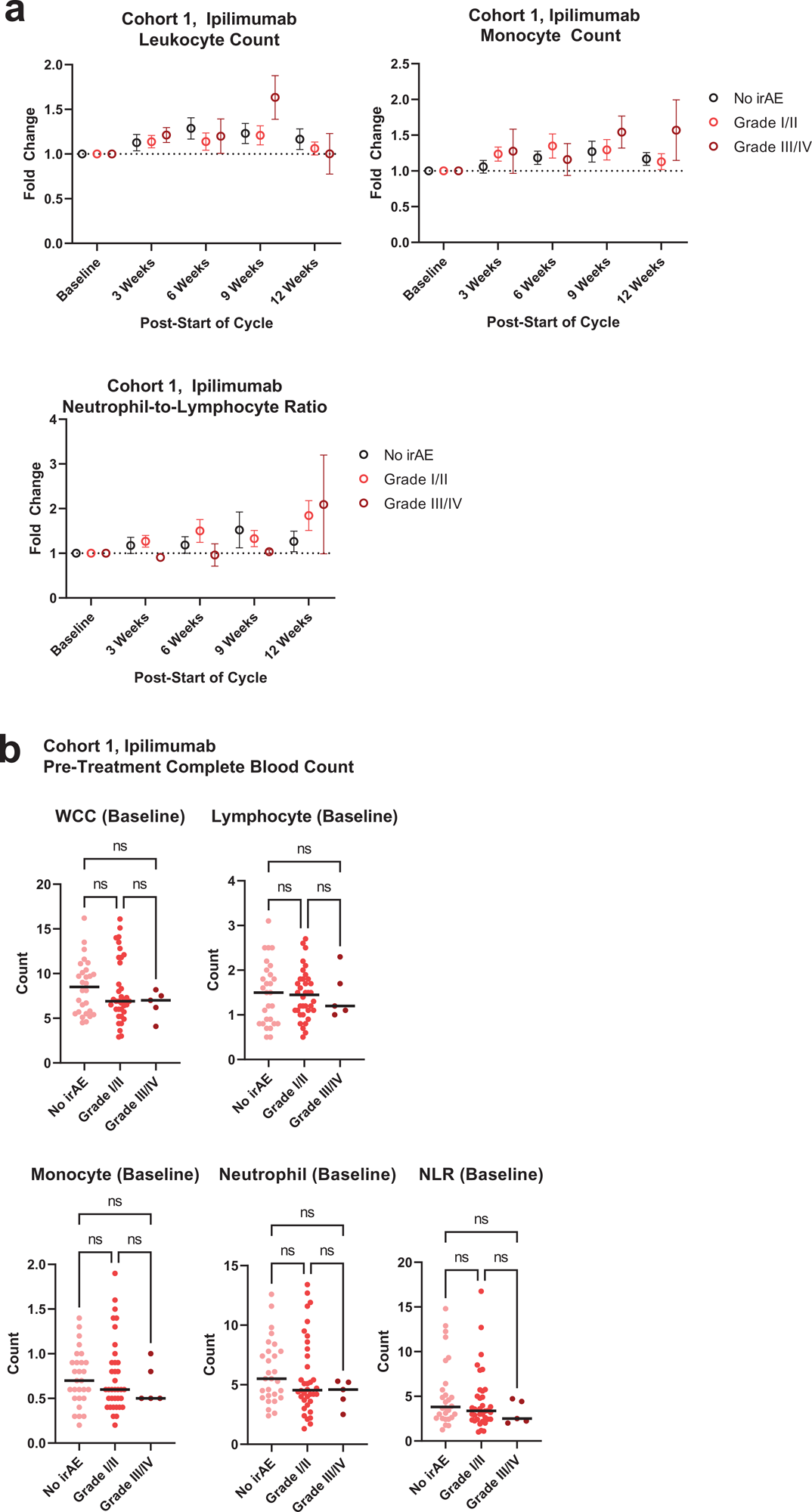
Immune cell numbers in the blood of ipilimumab-treated melanoma patients. (**a**) Lymphocyte, monocyte, and neutrophil-to-lymphocyte ratio fold changes by blood analyzer in cohort 1 ipilimumab-treated patients. Error = SEM (**b**) Immune cell counts by blood analyzed in cohort 1 baseline sampling. Bar = mean.

**Extended Data 5.**
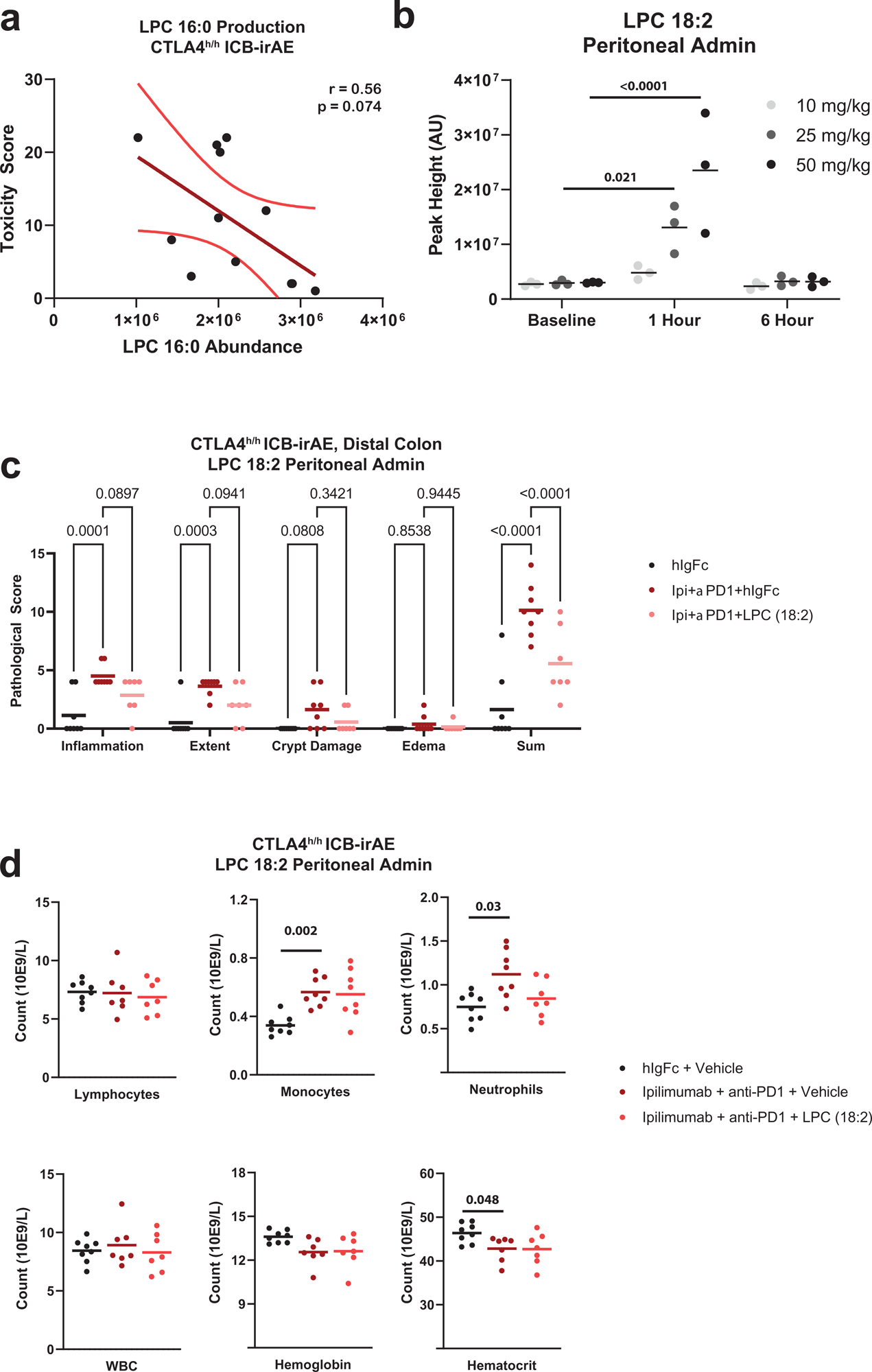
LPC loss and supplementation in *CTLA-4^h/h^* mice. (**a**) Scatter plot of composite toxicity score in *CTLA-4^h/h^* mice with matched LPC (16:0) concentration in terminal blood. Error = 95% CI. n = 12 (**b**) Blood plasma LPC 18:2 concentration following single dose intraperitoneal supplementation in C57BL/6 mice. Bar = mean. n = 3 per group. (**c**) Toxicity scoring by four criteria in *CTLA-4^h/h^* mice. Bar = mean. n = 7-8 per group. (**d**) Blood analysis by complete blood count in Day 30 *CTLA-4^h/h^* mice. Bar = mean. n = 7-8 per group. One-way ANOVA with Dunnett’s multiple comparisons test (**Extended Data 5b, 5d**); two-way ANOVA with Tukey’s multiple comparisons test (**Extended Data 5c**).

**Extended Data 6.**
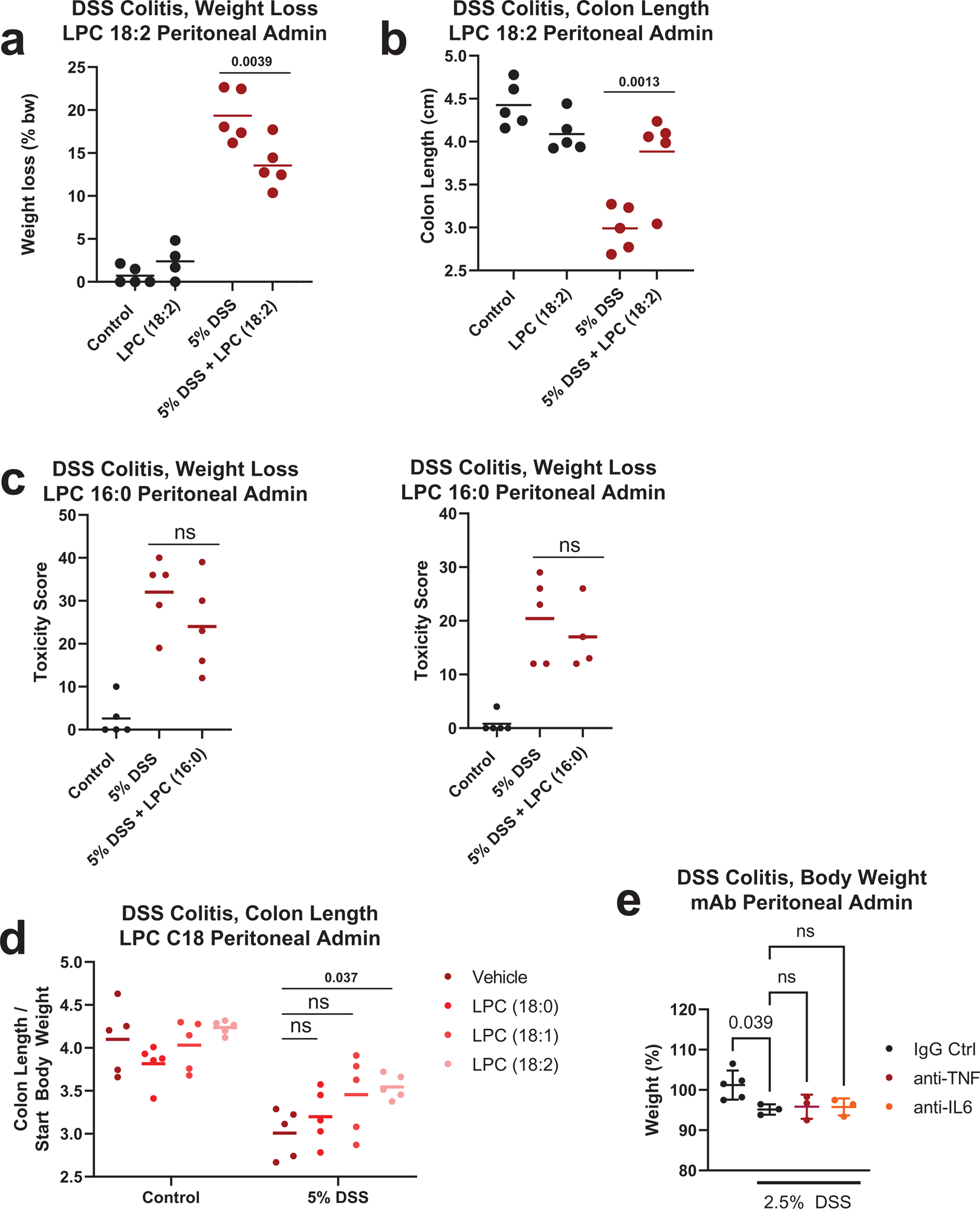
Loss and supplementation of LPC in DSS colitis mice. (**a-b**) Percent weight loss (**a**) and colon length relative to starting body weight (**b**) in 5% DSS treated mice following LPC 18:2 supplementation. N = 5 per group. Bar = mean. (**c**) Toxicity score in cecum and distal colon of 5% DSS treated mice following LPC (16:0) supplementation. n = 5 per group. Bar = mean. (**d**) Colon length relative to starting body weight in 5% DSS treated mice following supplementation with saturated and unsaturated C18-LPCs. n = 5 per group. Bar = mean. (**e**) Percent baseline weight in C57BL/6 mice receiving 2.5% DSS treated with 100ug control IgG, 100ug anti-TNF, or 100ug anti-IL-6 for 6 hours. n = 3-5 per group, Error = SD. Bar = mean. One-way ANOVA with Dunnett’s multiple comparisons test (**Extended Data 6a**); two-way ANOVA with Sidak’s multiple comparisons test (**Extended Data 6d-e**).

**Extended Data 7.**
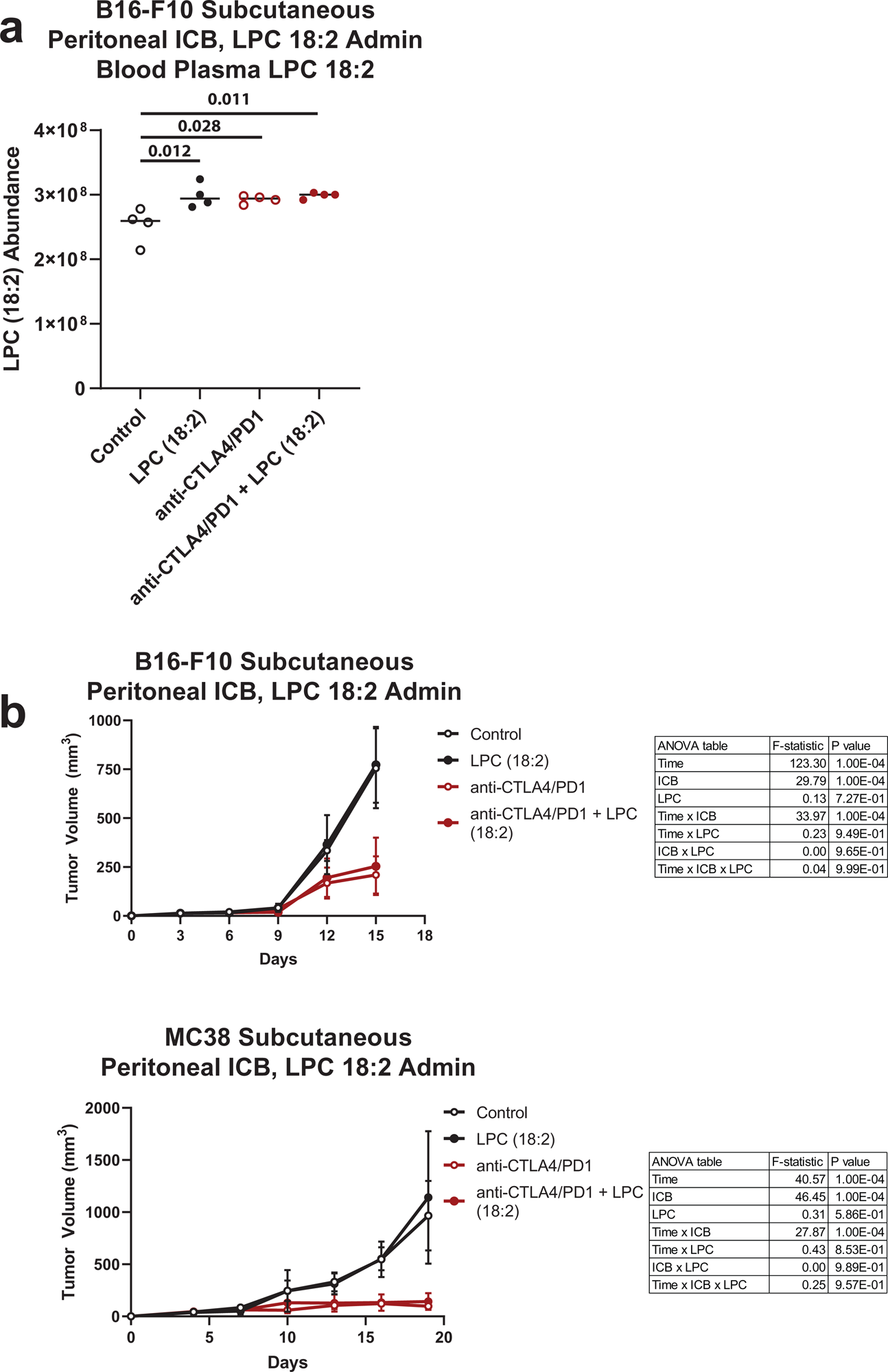
LPC 18:2 supplementation and ICB response in syngeneic tumor models. (**a**) LPC 18:2 abundance in terminal blood plasma from C57BL/6 mice bearing B16-F10 subcutaneous melanoma. n = 4 per group. Bar = mean. (**b**) B16-F10 or MC38 tumor volume following LPC 18:2 supplementation and anti-mouse CTLA-4 + anti-mouse PD-1 intraperitoneal treatment. n = 5 per group. Bar = mean. One-way ANOVA with Dunnett’s multiple comparisons test (**Extended Data 7a**); two-way ANOVA with Tukey’s multiple comparisons test (**Extended Data 7b**).

**Extended Data 8.**
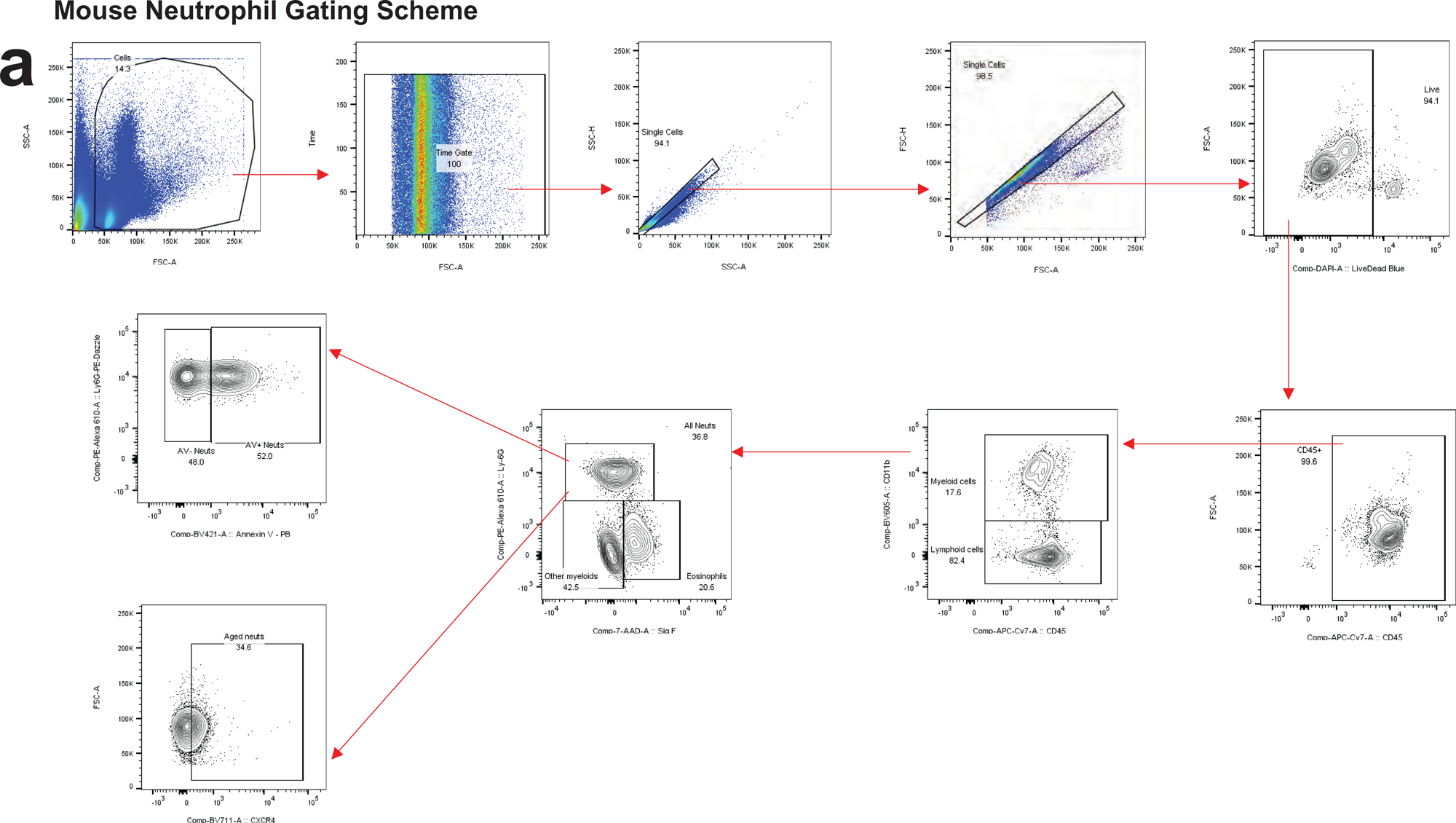
Flow cytometry gating scheme for neutrophils. **(a)** Strategy for gating mouse neutrophils (CD45^+^, CD11b^+^, Ly6G^+^).

## Notes

### Author Declarations

The North West-Greater Manchester Central Research Ethics Committee of Southampton University Hospitals NHS Foundation Trust gave ethical approval for the work included in Cohort 1 ("Southampton Ipilimumab") The University of California San Diego Human Research Protections Program Institutional Review Board of UC San Diego and Moores Cancer Center gave ethical approval for the work included in Cohort 2 ("Moores Cancer Center Mixed ICB") The Institutional Review Board of Yale University gave ethical approval for the work included in Cohort 3 ("Yale Pembrolizumab", NCT02407171) The Ethical Committee of Radboud University Nijmegen gave ethical approval for the work included from the Human Functional Genomics Project ("500FG") Coordinating Ethical Committee of the Helsinki and Uusimaa Hospital District gave ethical approval for the work included from the Finland National FINRISK study ("FINRISK 2002")

