## Supplementary Tables for "Linoleoyl-lysophosphatidylcholine suppresses immune-related adverse events due to immune checkpoint blockade"

### Supplementary Table 1

| | Cohort 1 (Ipilimumab) | Cohort 2 (Mixed ICB) | Cohort 3 (Pembrolizumab) |
|---|---|---|---|
| Location | UK Southampton | UCSD-Moores Cancer Center | Yale University |
| Total Patients (n) | 87 | 62 | 58 |
| Patients with Serial Sampling (n) | 65 | 32 | 53 |
| Total Serial Samples | 253 | 143 | 357 |
| Age (Range) | 61 (19-81) | 60 (27-87) | 68 (47-92) |
| Men (%) | 47 (54) | 36 (58) | 23 (40) |
| Tumor Histology (%) | Melanoma (100) | Melanoma (27), NSCLC (23), HNSCC (18), Breast (11), Colorectal (8), Skin (Non-Melanoma) (5), Renal (2), SCLC (2), Myoepithelial (2), Appendiceal (2), Small Bowel (2) | NSCLC (92), Melanoma (8) |
| Therapy (%) | Ipilimumab (100) | Pembrolizumab (42), Nivolumab (24), Ipilimumab + Nivolumab (21), Ipilimumab (6), Pembrolizumab + SBRT (3), Tremelimumab + anti-PD-L1 (2), Avelumab (2) | Pembrolizumab (78), Pembrolizumab + SBRT (22) |
| RECIST 1.1 % (CR:PR:SD:PD) | 1:11:10:60 | 0:13:27:48 | 0:30:16:42 |
| irAE Severity % (None : Grade I/II : Grade III/IV) | 41:52:7 | 52:31:18 | 28:61:12 |

### Supplementary Table 2

##### Cohort 1: Ipilimumab-Treated Advanced Melanoma
##### UK Southampton

| | Total | None | Grade I/II | Grade III/IV | p-value |
|---|---|---|---|---|---|
| n (%) | 65 (100) | 23 (35) | 37 (57) | 5 (8) | -- |
| Male (% per Group) | 37 (57) | 18 (78) | 18 (49) | 1 (20) | 0.11 |
| Age (Range) | 63 (19-81) | 57 (44-72) | 65 (19-76) | 70 (53-81) | 0.13 |
| Response (% per Group) | 17 (31) | 6 (40) | 10 (28) | 1 (33) | 0.83 |

### Supplementary Table 3

#### Cohort 1: Ipilimumab-Treated Melanoma
#### Maximum Excursion Associations for Identified Metabolites

| ID | class | adduct | m/z | irAE log(OR) | irAE pval | RECIST log(OR) | RECIST pval |
|---|---|---|---|---|---|---|---|
| b-Muricholic Acid | Bile Acid | [M-H] | 407.2805 | 0.08 [0.01 - 0.17] | 4.14E-02 | 0.02 [-0.07 - 0.12] | 7.00E-01 |
| Chenodeoxycholic Acid | Bile Acid | [M-H+Acetate] | 451.3068 | 0.05 [-0.03 - 0.13] | 2.74E-01 | -0.01 [-0.10 - 0.08] | 8.18E-01 |
| Deoxycholic Acid | Bile Acid | [M-H] | 391.2861 | -0.01 [-0.08 - 0.07] | 8.52E-01 | -0.03 [-0.12 - 0.05] | 3.99E-01 |
| Glycochenodeoxycholic Acid | Bile Acid | [M-H] | 448.3075 | -0.19 [-0.38 - -0.01] | 4.29E-02 | 0.04 [-0.13 - 0.22] | 6.46E-01 |
| Glycoursodeoxycholic Acid | Bile Acid | [M-H] | 448.3071 | -0.18 [-0.32 - -0.06] | 7.30E-03 | -0.08 [-0.22 - 0.05] | 2.45E-01 |
| Lithocholic Acid | Bile Acid | [M-H+Acetate] | 435.3115 | -0.08 [-0.22 - 0.06] | 2.76E-01 | -0.05 [-0.20 - 0.10] | 4.83E-01 |
| Taurochenodeoxycholic Acid | Bile Acid | [M-H] | 498.2843 | -0.02 [-0.16 - 0.12] | 7.82E-01 | -0.10 [-0.25 - 0.04] | 1.59E-01 |
| Tauroursodeoxycholic Acid | Bile Acid | [M-H] | 498.2903 | -0.14 [-0.28 - -0.02] | 3.20E-02 | -0.04 [-0.17 - 0.09] | 5.92E-01 |
| 10,11-DiHDPA | Docosanoid | [M-H] | 361.2386 | 0.21 [-0.02 - 0.46] | 7.93E-02 | 0.03 [-0.22 - 0.27] | 8.20E-01 |
| 13,14-EpDPA | Docosanoid | [M-H] | 343.2283 | 0.06 [-0.08 - 0.20] | 4.36E-01 | 0.02 [-0.13 - 0.19] | 7.55E-01 |
| 19,20-DiHDPA | Docosanoid | [M-H] | 361.2389 | -0.13 [-0.42 - 0.14] | 3.51E-01 | 0.13 [-0.14 - 0.43] | 3.58E-01 |
| 7,8-DiHDPA | Docosanoid | [M-H] | 361.2423 | -1.19 [-2.49 - 0.04] | 6.39E-02 | 0.41 [-0.80 - 1.68] | 5.12E-01 |
| MCTR2 | Docosanoid | [M-H] | 519.247 | 0.04 [-0.29 - 0.38] | 8.18E-01 | 0.02 [-0.33 - 0.39] | 9.00E-01 |
| Resolvin D1 | Docosanoid | [M-H] | 375.2218 | 0.09 [-0.18 - 0.37] | 5.08E-01 | 0.14 [-0.16 - 0.47] | 3.86E-01 |
| 11(12)-EpETE | Eicosanoid | [M-H] | 317.2125 | 0.22 [0.04 - 0.43] | 2.42E-02 | 0.26 [0.05 - 0.51] | 2.14E-02 |
| 11,12-diHETrE | Eicosanoid | [M-H+Acetate] | 397.2578 | 0.09 [-0.15 - 0.33] | 4.57E-01 | 0.11 [-0.16 - 0.42] | 4.51E-01 |
| 11b dhk PGF2a | Eicosanoid | [M-H] | 353.2338 | 0.12 [-0.31 - 0.55] | 5.88E-01 | 0.13 [-0.34 - 0.62] | 5.97E-01 |
| 11-beta-PGE1 | Eicosanoid | [M-H] | 353.2336 | -0.04 [-0.31 - 0.22] | 7.39E-01 | 0.07 [-0.23 - 0.37] | 6.41E-01 |
| 11-dehydro-2,3-dinor-TXB2 | Eicosanoid | [M-H+Acetate] | 399.1949 | 0.06 [-0.09 - 0.21] | 4.65E-01 | -0.03 [-0.22 - 0.15] | 7.77E-01 |
| 11-HETE | Eicosanoid | [M-H] | 319.2281 | 0.12 [0.01 - 0.23] | 4.12E-02 | 0.11 [-0.01 - 0.23] | 8.10E-02 |
| 12,13 diHOME | Eicosanoid | [M-H] | 313.2388 | 0.09 [-0.06 - 0.25] | 2.52E-01 | -0.02 [-0.19 - 0.15] | 7.95E-01 |
| 12,13 EpOME | Eicosanoid | [M-H] | 295.2281 | 0.03 [-0.27 - 0.34] | 8.34E-01 | 0.17 [-0.16 - 0.53] | 3.17E-01 |
| 12-HHTrE | Eicosanoid | [M-H] | 279.1969 | 0.08 [0.01 - 0.15] | 3.41E-02 | 0.12 [0.03 - 0.23] | 1.60E-02 |
| 12-OPDA | Eicosanoid | [M-H+Acetate] | 351.2162 | 0.01 [-0.28 - 0.29] | 9.65E-01 | 0.25 [-0.08 - 0.63] | 1.53E-01 |
| 12-oxoETE | Eicosanoid | [M-H] | 317.2122 | -0.01 [-0.20 - 0.17] | 8.97E-01 | 0.07 [-0.13 - 0.27] | 4.73E-01 |
| 12S-HpEPE | Eicosanoid | [M-H] | 333.205 | -0.08 [-0.28 - 0.12] | 4.51E-01 | 0.33 [0.05 - 0.65] | 3.26E-02 |

| ID | class | adduct | m/z | irAE log(OR) | irAE pval | RECIST log(OR) | RECIST pval |
|---|---|---|---|---|---|---|---|
| 12S-HpETE | Eicosanoid | [M-H] | 335.2228 | 0.33 [0.07 - 0.63] | 1.77E-02 | 0.28 [0.00 - 0.58] | 5.16E-02 |
| 13(S) HOTrE(y) | Eicosanoid | [M-H+Acetate] | 353.2313 | 0.08 [-0.19 - 0.35] | 5.45E-01 | 0.36 [0.05 - 0.72] | 3.33E-02 |
| 13,14-dihydro-15-keto-PGA2 | Eicosanoid | [M-H+Acetate] | 393.2287 | -0.03 [-0.19 - 0.13] | 7.56E-01 | -0.08 [-0.28 - 0.10] | 3.87E-01 |
| 13,14-dihydro-15-keto-tetranor-PGF1alpha | Eicosanoid | [M-H+Acetate] | 359.2108 | -0.02 [-0.58 - 0.55] | 9.50E-01 | -0.07 [-0.68 - 0.54] | 8.12E-01 |
| 13,14-dihydro-PGE1 | Eicosanoid | [M-H] | 355.2462 | -0.10 [-0.87 - 0.69] | 8.09E-01 | 0.39 [-0.53 - 1.73] | 5.04E-01 |
| 13-HODE | Eicosanoid | [M-H] | 295.228 | -0.08 [-0.27 - 0.10] | 4.15E-01 | 0.10 [-0.09 - 0.30] | 3.11E-01 |
| 13-HpODE | Eicosanoid | [M-H] | 311.2233 | -0.11 [-0.37 - 0.14] | 3.80E-01 | 0.05 [-0.25 - 0.34] | 7.57E-01 |
| 13-oxoODE | Eicosanoid | [M-H+Acetate] | 353.2315 | -0.05 [-0.30 - 0.18] | 6.71E-01 | 0.12 [-0.14 - 0.42] | 3.68E-01 |
| 13S-HpOTrEγ | Eicosanoid | [M-H] | 309.2074 | -0.10 [-0.31 - 0.11] | 3.69E-01 | 0.14 [-0.12 - 0.42] | 3.15E-01 |
| 14(15) EpETE | Eicosanoid | [M-H] | 317.2119 | 0.20 [-0.11 - 0.53] | 2.18E-01 | NA [NA - NA] | NA |
| 14,15 LTD4 | Eicosanoid | [M-H] | 495.2607 | 0.15 [-0.04 - 0.36] | 1.26E-01 | 0.14 [-0.08 - 0.38] | 2.26E-01 |
| 14,15-DiHETE | Eicosanoid | [M-H+Acetate] | 395.2419 | 0.04 [-0.18 - 0.27] | 7.25E-01 | -0.18 [-0.47 - 0.07] | 1.81E-01 |
| 14,15-diHETrE | Eicosanoid | [M-H+Acetate] | 397.2624 | 0.40 [0.01 - 0.83] | 5.66E-02 | 0.22 [-0.22 - 0.68] | 3.36E-01 |
| 14,15-EET | Eicosanoid | [M-H] | 319.2292 | 0.02 [-0.18 - 0.23] | 8.25E-01 | 0.09 [-0.13 - 0.32] | 4.06E-01 |
| 15 oxoEDE | Eicosanoid | [M-H] | 321.2437 | 0.06 [-0.20 - 0.33] | 6.35E-01 | 0.19 [-0.08 - 0.48] | 1.82E-01 |
| 15(S) HEPE | Eicosanoid | [M-H] | 317.2124 | 0.03 [-0.08 - 0.14] | 5.75E-01 | 0.02 [-0.10 - 0.14] | 6.96E-01 |
| 15(S) HETrE | Eicosanoid | [M-H] | 321.2437 | 0.04 [-0.06 - 0.14] | 4.07E-01 | 0.06 [-0.05 - 0.17] | 2.60E-01 |
| 15d PGA2 | Eicosanoid | [M-H] | 315.1967 | 0.23 [-0.14 - 0.66] | 2.49E-01 | -0.16 [-0.65 - 0.25] | 4.73E-01 |
| 15d PGJ2 | Eicosanoid | [M-H] | 315.2003 | 0.11 [-0.24 - 0.48] | 5.40E-01 | 0.32 [-0.08 - 0.75] | 1.30E-01 |
| 15-epi-PGA1 | Eicosanoid | [M-H+Acetate] | 395.2441 | -0.02 [-0.60 - 0.55] | 9.34E-01 | -0.12 [-0.72 - 0.45] | 6.94E-01 |
| 15-HETE | Eicosanoid | [M-H+Acetate] | 379.2473 | 0.09 [-0.19 - 0.36] | 5.27E-01 | 0.04 [-0.25 - 0.34] | 7.92E-01 |
| 15-keto-PGA1 | Eicosanoid | [M-H+Acetate] | 393.2307 | 0.19 [-0.14 - 0.54] | 2.72E-01 | -0.10 [-0.48 - 0.26] | 5.96E-01 |
| 15S-HpETE | Eicosanoid | [M-H] | 335.2227 | 0.09 [-0.14 - 0.33] | 4.44E-01 | 0.32 [0.03 - 0.64] | 3.93E-02 |
| 17k DPA | Eicosanoid | [M-H+Acetate] | 403.2462 | 0.08 [-0.04 - 0.21] | 1.91E-01 | NA [NA - NA] | NA |
| 18(+/-)-HETE | Eicosanoid | [M-H] | 319.2232 | 0.15 [-0.47 - 0.77] | 6.40E-01 | 0.73 [-0.08 - 1.71] | 1.06E-01 |
| 20cooh AA | Eicosanoid | [M-H] | 333.2083 | -0.13 [-0.62 - 0.36] | 5.96E-01 | 0.05 [-0.49 - 0.58] | 8.57E-01 |
| 5(S) HEPE | Eicosanoid | [M-H+Acetate] | 377.2312 | 0.04 [-0.22 - 0.29] | 7.71E-01 | 0.05 [-0.22 - 0.32] | 7.07E-01 |

| ID | class | adduct | m/z | irAE log(OR) | irAE pval | RECIST log(OR) | RECIST pval |
|---|---|---|---|---|---|---|---|
| 5(S) HETrE | Eicosanoid | [M-H] | 321.2437 | -0.07 [-0.29 - 0.15] | 5.44E-01 | 0.21 [-0.01 - 0.45] | 7.17E-02 |
| 5,12-DiHETE | Eicosanoid | [M-H] | 335.2232 | 0.12 [-0.03 - 0.28] | 1.26E-01 | 0.13 [-0.03 - 0.30] | 1.16E-01 |
| 5,6-diHETE | Eicosanoid | [M-H] | 335.2238 | -0.04 [-0.38 - 0.29] | 7.89E-01 | 0.22 [-0.14 - 0.61] | 2.39E-01 |
| 5,6-diHETrE | Eicosanoid | [M-H] | 337.2386 | 0.07 [-0.36 - 0.49] | 7.59E-01 | -0.16 [-0.66 - 0.33] | 5.33E-01 |
| 5,6-EET | Eicosanoid | [M-H] | 319.2281 | 0.09 [-0.14 - 0.32] | 4.17E-01 | 0.20 [-0.05 - 0.45] | 1.16E-01 |
| 5-HETE | Eicosanoid | [M-H] | 319.2282 | 0.09 [-0.02 - 0.22] | 1.19E-01 | 0.14 [0.01 - 0.29] | 4.57E-02 |
| 5-oxoETE | Eicosanoid | [M-H] | 317.2126 | 0.00 [-0.21 - 0.20] | 9.83E-01 | 0.11 [-0.07 - 0.30] | 2.36E-01 |
| 5S-HpEPE | Eicosanoid | [M-H] | 333.205 | -0.14 [-0.40 - 0.10] | 2.54E-01 | 0.45 [0.16 - 0.79] | 4.95E-03 |
| 6t LTB4 | Eicosanoid | [M-H] | 335.2232 | 0.13 [-0.01 - 0.29] | 8.19E-02 | 0.15 [-0.01 - 0.33] | 7.90E-02 |
| 7 HDoHE | Eicosanoid | [M-H] | 343.2277 | 0.03 [-0.14 - 0.20] | 7.39E-01 | 0.06 [-0.13 - 0.26] | 5.32E-01 |
| 8(9)-EpETE | Eicosanoid | [M-H+Acetate] | 377.2303 | -0.06 [-0.31 - 0.20] | 6.61E-01 | -0.03 [-0.30 - 0.24] | 8.08E-01 |
| 8(S) HEPE | Eicosanoid | [M-H] | 317.2125 | 0.11 [-0.06 - 0.29] | 2.00E-01 | 0.07 [-0.12 - 0.27] | 4.45E-01 |
| 8(S) HETrE | Eicosanoid | [M-H+Acetate] | 381.2614 | -0.14 [-0.36 - 0.05] | 1.71E-01 | 0.09 [-0.11 - 0.29] | 3.60E-01 |
| 8,12-iso-iPF2à-VI-1,5-lactone | Eicosanoid | [M-H] | 335.2229 | 0.15 [-0.11 - 0.42] | 2.60E-01 | 0.21 [-0.10 - 0.53] | 1.90E-01 |
| 8,12-iso-iPF2à-VIÿ | Eicosanoid | [M-H] | 353.2334 | 0.20 [0.02 - 0.39] | 3.64E-02 | 0.12 [-0.07 - 0.31] | 2.08E-01 |
| 8,15-diHETE | Eicosanoid | [M-H] | 335.2231 | 0.17 [-0.11 - 0.45] | 2.28E-01 | 0.15 [-0.16 - 0.48] | 3.35E-01 |
| 8,9-diHETrE | Eicosanoid | [M-H+Acetate] | 397.2574 | 0.03 [-0.16 - 0.23] | 7.29E-01 | 0.09 [-0.13 - 0.31] | 4.25E-01 |
| 8,9-EET | Eicosanoid | [M-H] | 319.2287 | 0.06 [-0.23 - 0.36] | 6.69E-01 | NA [NA - NA] | NA |
| 8-iso-PGA1 | Eicosanoid | [M-H] | 335.2231 | -0.09 [-0.31 - 0.12] | 3.85E-01 | -0.07 [-0.33 - 0.17] | 5.61E-01 |
| 8-iso-PGF1a | Eicosanoid | [M-H] | 355.2485 | -0.27 [-0.55 - -0.02] | 4.39E-02 | -0.03 [-0.32 - 0.25] | 8.11E-01 |
| 9(S) HOTrE | Eicosanoid | [M-H] | 293.2126 | 0.08 [-0.08 - 0.26] | 3.21E-01 | 0.07 [-0.11 - 0.25] | 4.69E-01 |
| 9,10 EpOME | Eicosanoid | [M-H] | 295.2269 | 0.10 [-0.36 - 0.56] | 6.64E-01 | 0.21 [-0.25 - 0.67] | 3.58E-01 |
| 9-HpODE | Eicosanoid | [M-H] | 311.2217 | -0.10 [-0.30 - 0.09] | 2.96E-01 | 0.04 [-0.19 - 0.25] | 6.92E-01 |
| 9-oxoODE | Eicosanoid | [M-H] | 293.2125 | -0.05 [-0.37 - 0.26] | 7.47E-01 | 0.02 [-0.30 - 0.35] | 8.86E-01 |
| 9-oxoOTrE | Eicosanoid | [M-H] | 291.1968 | 0.04 [-0.16 - 0.25] | 6.79E-01 | 0.10 [-0.11 - 0.34] | 3.51E-01 |
| 9S-HpOTrE | Eicosanoid | [M-H] | 309.2074 | -0.03 [-0.27 - 0.21] | 7.90E-01 | 0.19 [-0.10 - 0.49] | 2.03E-01 |
| ent-8-iso Prostaglandin F2α | Eicosanoid | [M-H] | 353.234 | -0.05 [-0.58 - 0.50] | 8.66E-01 | 0.66 [-0.11 - 1.64] | 1.50E-01 |

| ID | class | adduct | m/z | irAE log(OR) | irAE pval | RECIST log(OR) | RECIST pval |
|---|---|---|---|---|---|---|---|
| HXA3 | Eicosanoid | [M-H] | 335.2234 | 0.20 [0.02 - 0.40] | 3.84E-02 | 0.20 [0.00 - 0.42] | 5.98E-02 |
| PDX | Eicosanoid | [M-H+Acetate] | 419.2386 | 0.35 [-0.12 - 0.87] | 1.58E-01 | 0.92 [0.35 - 1.60] | 3.15E-03 |
| PGD2 | Eicosanoid | [M-H] | 351.218 | 0.09 [-0.12 - 0.30] | 4.25E-01 | -0.02 [-0.25 - 0.19] | 8.24E-01 |
| Resolvin E1 | Eicosanoid | [M-H] | 349.2023 | 0.04 [-0.05 - 0.13] | 4.15E-01 | -0.03 [-0.13 - 0.07] | 5.14E-01 |
| tetranor 12(R) HETE | Eicosanoid | [M-H+Acetate] | 325.2027 | -0.06 [-0.29 - 0.17] | 6.30E-01 | 0.69 [0.32 - 1.17] | 1.36E-03 |
| D-erythro-Sphingosine C-18 | Endocannabinoid | [M-H+Acetate] | 358.2961 | 0.22 [-0.15 - 0.61] | 2.53E-01 | 0.38 [-0.06 - 0.85] | 9.88E-02 |
| Docosahexaenoyl Ethanolamide | Endocannabinoid | [M-H+Acetate] | 430.2968 | 0.20 [-0.13 - 0.55] | 2.38E-01 | 0.26 [-0.08 - 0.62] | 1.42E-01 |
| N-oleoyl Dopamine | Endocannabinoid | [M-H] | 416.3105 | 0.12 [-0.08 - 0.33] | 2.40E-01 | 0.04 [-0.17 - 0.27] | 6.84E-01 |
| Oleoyl Ethanolamide | Endocannabinoid | [M-H+Acetate] | 384.3121 | 0.00 [-0.28 - 0.28] | 9.99E-01 | -0.03 [-0.35 - 0.28] | 8.36E-01 |
| Adrenic Acid | Free Fatty Acid | [M-H] | 331.2603 | -0.02 [-0.15 - 0.12] | 7.90E-01 | 0.03 [-0.14 - 0.19] | 7.05E-01 |
| a-linolenic Acid | Free Fatty Acid | [M-H] | 277.2172 | 0.04 [-0.14 - 0.23] | 6.38E-01 | 0.13 [-0.07 - 0.34] | 2.18E-01 |
| Arachidic Acid | Free Fatty Acid | [M-H] | 311.2957 | 0.11 [-0.30 - 0.53] | 6.08E-01 | 0.56 [0.07 - 1.12] | 3.47E-02 |
| Arachidonic Acid | Free Fatty Acid | [M-H] | 303.2329 | 0.17 [-0.20 - 0.54] | 3.66E-01 | 0.00 [-0.44 - 0.41] | 9.89E-01 |
| Behenic Acid | Free Fatty Acid | [M-H] | 339.3269 | 0.09 [-0.15 - 0.34] | 4.51E-01 | 0.10 [-0.16 - 0.39] | 4.62E-01 |
| Dihomo-g-linolenic Acid | Free Fatty Acid | [M-H] | 305.2492 | 0.19 [-0.11 - 0.51] | 2.32E-01 | 0.05 [-0.29 - 0.40] | 7.75E-01 |
| Docosadienoic Acid | Free Fatty Acid | [M-H] | 335.2961 | 0.04 [-0.18 - 0.26] | 7.53E-01 | 0.13 [-0.13 - 0.38] | 3.29E-01 |
| Docosaenoic Acid | Free Fatty Acid | [M-H] | 337.3118 | 0.14 [-0.11 - 0.39] | 2.78E-01 | 0.10 [-0.16 - 0.37] | 4.47E-01 |
| Docosahexaenoic Acid | Free Fatty Acid | [M-H] | 327.2329 | -0.02 [-0.30 - 0.25] | 8.65E-01 | 0.20 [-0.10 - 0.52] | 1.94E-01 |
| Docosapentaenoic Acid | Free Fatty Acid | [M-H] | 329.2491 | 0.01 [-0.17 - 0.19] | 9.19E-01 | 0.11 [-0.09 - 0.32] | 2.74E-01 |
| Docosatrienoic Acid | Free Fatty Acid | [M-H] | 333.2805 | 0.04 [-0.15 - 0.24] | 6.55E-01 | 0.10 [-0.10 - 0.32] | 3.24E-01 |
| Eicosadienoic Acid | Free Fatty Acid | [M-H] | 307.2645 | 0.01 [-0.18 - 0.20] | 9.43E-01 | 0.22 [0.01 - 0.45] | 4.60E-02 |
| Eicosapentaenoic Acid | Free Fatty Acid | [M-H] | 301.2173 | 0.03 [-0.13 - 0.20] | 7.06E-01 | 0.02 [-0.15 - 0.29] | 8.10E-01 |
| Eicosatrienoic Acid | Free Fatty Acid | [M-H] | 305.2488 | 0.10 [-0.12 - 0.34] | 3.68E-01 | 0.08 [-0.18 - 0.34] | 5.48E-01 |
| Eicosenoic Acid | Free Fatty Acid | [M-H] | 309.2799 | 0.05 [-0.11 - 0.22] | 5.29E-01 | 0.16 [-0.02 - 0.35] | 8.05E-02 |
| Gondolic Acid | Free Fatty Acid | [M-H] | 309.2799 | 0.04 [-0.12 - 0.21] | 5.91E-01 | 0.16 [-0.02 - 0.35] | 7.55E-02 |
| Heptadecaenoic Acid | Free Fatty Acid | [M-H+Acetate] | 327.2545 | -0.07 [-0.18 - 0.05] | 2.64E-01 | -0.04 [-0.17 - 0.08] | 5.36E-01 |
| Heptadecanoic Acid | Free Fatty Acid | [M-H] | 269.2485 | 0.03 [-0.26 - 0.31] | 8.62E-01 | 0.25 [-0.07 - 0.59] | 1.43E-01 |

| ID | class | adduct | m/z | irAE log(OR) | irAE pval | RECIST log(OR) | RECIST pval |
|---|---|---|---|---|---|---|---|
| Lignoceric Acid | Free Fatty Acid | [M-H] | 367.3581 | 0.12 [-0.17 - 0.41] | 4.27E-01 | 0.28 [-0.08 - 0.74] | 1.76E-01 |
| Linoleic Acid | Free Fatty Acid | [M-H] | 279.2341 | 0.02 [-0.15 - 0.18] | 8.41E-01 | 0.06 [-0.12 - 0.23] | 5.26E-01 |
| Myristic Acid | Free Fatty Acid | [M-H] | 227.2013 | 0.00 [-0.54 - 0.55] | 9.87E-01 | -0.39 [-0.98 - 0.19] | 1.85E-01 |
| Nervonic Acid | Free Fatty Acid | [M-H] | 365.3427 | 0.10 [-0.23 - 0.45] | 5.45E-01 | 0.36 [-0.01 - 0.76] | 6.32E-02 |
| Oleic Acid | Free Fatty Acid | [M-H] | 281.2487 | 0.00 [-0.17 - 0.17] | 9.72E-01 | 0.08 [-0.12 - 0.27] | 4.38E-01 |
| Osbond Acid | Free Fatty Acid | [M-H] | 329.248 | 0.04 [-0.14 - 0.22] | 6.62E-01 | 0.14 [-0.06 - 0.34] | 1.67E-01 |
| Palmitic Acid | Free Fatty Acid | [M-H+Acetate] | 315.2544 | -0.06 [-0.30 - 0.16] | 5.88E-01 | 0.23 [-0.03 - 0.51] | 8.64E-02 |
| Palmitoleic Acid | Free Fatty Acid | [M-H+Acetate] | 313.2388 | -0.04 [-0.27 - 0.18] | 7.06E-01 | -0.16 [-0.42 - 0.09] | 2.10E-01 |
| Pentadecanoic Acid | Free Fatty Acid | [M-H] | 241.2172 | 0.09 [-0.27 - 0.46] | 6.21E-01 | 0.25 [-0.13 - 0.67] | 2.05E-01 |
| Petroselenic Acid | Free Fatty Acid | [M-H] | 341.2701 | 0.01 [-0.19 - 0.20] | 9.45E-01 | 0.05 [-0.18 - 0.28] | 6.93E-01 |
| Tricosanoic Acid | Free Fatty Acid | [M-H] | 353.3427 | -0.22 [-0.50 - 0.05] | 1.22E-01 | -0.02 [-0.31 - 0.28] | 8.89E-01 |
| Tricosenoic Acid | Free Fatty Acid | [M-H] | 351.3271 | -0.13 [-0.46 - 0.19] | 4.37E-01 | 0.10 [-0.23 - 0.43] | 5.64E-01 |
| LPC (14:0), Peak 1 | Lysolipid | [M+Cl-] | 502.2706 | -0.22 [-0.76 - 0.30] | 4.02E-01 | -1.01 [-1.73 - -0.38] | 3.20E-03 |
| LPC (14:0), Peak 2 | Lysolipid | [M+Cl-] | 502.2713 | 0.08 [-0.48 - 0.65] | 7.84E-01 | -0.75 [-1.52 - -0.09] | 3.73E-02 |
| LPC (15:0), Peak 1 | Lysolipid | [M-H+Acetate] | 540.3315 | 0.14 [-0.49 - 0.78] | 6.67E-01 | -1.36 [-2.32 - -0.54] | 2.41E-03 |
| LPC (15:0), Peak 2 | Lysolipid | [M-H+Acetate] | 540.3314 | -0.02 [-0.65 - 0.63] | 9.61E-01 | -0.78 [-1.89 - -0.01] | 9.71E-02 |
| LPC (16:0), Peak 1 | Lysolipid | [M+Cl-] | 530.3024 | -1.03 [-4.72 - 2.46] | 5.60E-01 | -2.90 [-7.83 - 1.28] | 2.12E-01 |
| LPC (16:0), Peak 2 | Lysolipid | [M+Cl-] | 530.3024 | -6.53 [-11.46 - -2.28] | 4.63E-03 | 4.68 [0.02 - 9.92] | 6.22E-02 |
| LPC (16:1), Peak 1 | Lysolipid | [M-H+Acetate] | 552.3334 | 0.20 [-0.44 - 0.86] | 5.40E-01 | -1.01 [-1.89 - -0.21] | 1.70E-02 |
| LPC (16:1), Peak 2 | Lysolipid | [M-H+Acetate] | 552.3314 | 0.17 [-0.28 - 0.62] | 4.67E-01 | -0.11 [-0.62 - 0.46] | 6.50E-01 |
| LPC (17:0), Peak 1 | Lysolipid | [M-H+Acetate] | 568.363 | -0.11 [-0.78 - 0.55] | 7.40E-01 | -1.40 [-2.42 - -0.51] | 3.64E-03 |
| LPC (17:0), Peak 2 | Lysolipid | [M-H+Acetate] | 568.3621 | -0.05 [-0.69 - 0.59] | 8.69E-01 | -0.63 [-1.59 - 0.10] | 1.26E-01 |
| LPC (17:1), Peak 1 | Lysolipid | [M-H+Acetate] | 566.3475 | 0.26 [-0.26 - 0.80] | 3.23E-01 | -0.33 [-1.02 - 0.26] | 2.71E-01 |
| LPC (17:1), Peak 2 | Lysolipid | [M-H+Acetate] | 566.3502 | 0.18 [-0.53 - 0.91] | 6.24E-01 | -1.58 [-2.74 - -0.59] | 3.50E-03 |
| LPC (18:0), Peak 1 | Lysolipid | [M+Cl-] | 558.3333 | -0.02 [-0.51 - 0.48] | 9.36E-01 | -0.07 [-0.62 - 0.63] | 7.91E-01 |
| LPC (18:0), Peak 2 | Lysolipid | [M+Cl-] | 558.3334 | -0.63 [-2.36 - 1.12] | 4.70E-01 | -0.49 [-2.40 - 1.44] | 6.05E-01 |
| LPC (18:1), Peak 1 | Lysolipid | [M-H+Acetate] | 580.3627 | 0.19 [-0.29 - 0.67] | 4.30E-01 | -0.11 [-0.66 - 0.50] | 6.70E-01 |

| ID | class | adduct | m/z | irAE log(OR) | irAE pval | RECIST log(OR) | RECIST pval |
|---|---|---|---|---|---|---|---|
| LPC (18:1), Peak 2 | Lysolipid | [M-H+Acetate] | 580.3619 | 0.53 [-0.84 - 1.91] | 4.48E-01 | -0.42 [-2.00 - 1.21] | 5.64E-01 |
| LPC (18:2), Peak 1 | Lysolipid | [M+Cl-] | 554.3004 | -0.15 [-1.15 - 0.85] | 7.60E-01 | -1.14 [-2.43 - 0.02] | 6.26E-02 |
| LPC (18:2), Peak 2 | Lysolipid | [M+Cl-] | 554.3024 | -4.53 [-9.35 - -0.21] | 4.74E-02 | 2.19 [-2.41 - 7.53] | 3.80E-01 |
| LPC (18:3), Peak 1 | Lysolipid | [M-H+Acetate] | 576.3323 | 0.03 [-0.43 - 0.49] | 9.09E-01 | -0.28 [-0.80 - 0.25] | 2.82E-01 |
| LPC (18:3), Peak 2 | Lysolipid | [M-H+Acetate] | 576.3314 | -0.04 [-0.48 - 0.39] | 8.44E-01 | -0.16 [-0.63 - 0.33] | 5.13E-01 |
| LPC (19:1) | Lysolipid | [M-H+Acetate] | 594.3799 | 0.34 [-0.23 - 0.92] | 2.46E-01 | -0.57 [-1.42 - 0.08] | 1.17E-01 |
| LPC (20:1), Peak 1 | Lysolipid | [M-H+Acetate] | 608.3909 | -0.35 [-1.20 - 0.48] | 4.11E-01 | -1.39 [-2.69 - -0.29] | 2.12E-02 |
| LPC (20:1), Peak 2 | Lysolipid | [M-H+Acetate] | 608.3928 | -0.20 [-0.94 - 0.54] | 5.95E-01 | -0.88 [-1.91 - 0.02] | 6.69E-02 |
| LPC (20:2), Peak 1 | Lysolipid | [M-H+Acetate] | 606.3781 | -0.01 [-0.55 - 0.55] | 9.77E-01 | -0.51 [-1.25 - 0.12] | 1.29E-01 |
| LPC (20:2), Peak 2 | Lysolipid | [M-H+Acetate] | 606.3787 | 0.22 [-0.34 - 0.79] | 4.36E-01 | -0.26 [-0.91 - 0.37] | 4.10E-01 |
| LPC (20:3), Peak 1 | Lysolipid | [M-H+Acetate] | 604.3636 | 0.28 [-0.41 - 1.00] | 4.29E-01 | -0.65 [-1.50 - 0.14] | 1.16E-01 |
| LPC (20:3), Peak 2 | Lysolipid | [M-H+Acetate] | 604.363 | 0.19 [-0.37 - 0.74] | 5.13E-01 | -0.28 [-0.99 - 0.34] | 3.58E-01 |
| LPC (20:4), Peak 1 | Lysolipid | [M-H+Acetate] | 602.3474 | 0.30 [-0.69 - 1.34] | 5.54E-01 | -0.75 [-2.01 - 0.41] | 2.17E-01 |
| LPC (20:4), Peak 2 | Lysolipid | [M-H+Acetate] | 602.3475 | 0.74 [-1.02 - 2.61] | 4.15E-01 | -1.88 [-4.37 - 0.28] | 1.10E-01 |
| LPC (20:5), Peak 1 | Lysolipid | [M-H+Acetate] | 600.3315 | 0.06 [-0.46 - 0.60] | 8.11E-01 | -0.55 [-1.25 - 0.10] | 1.08E-01 |
| LPC (20:5), Peak 2 | Lysolipid | [M-H+Acetate] | 600.3321 | 0.02 [-0.35 - 0.40] | 9.02E-01 | -0.05 [-0.47 - 0.46] | 8.10E-01 |
| LPC (22:4), Peak 2 | Lysolipid | [M-H+Acetate] | 630.3779 | 0.51 [-0.16 - 1.20] | 1.36E-01 | -0.11 [-0.85 - 0.69] | 7.72E-01 |
| LPE (14:0), Peak 1 | Lysolipid | [M-H] | 424.2472 | -0.02 [-0.36 - 0.32] | 9.11E-01 | -0.44 [-0.88 - -0.05] | 3.44E-02 |
| LPE (14:0), Peak 2 | Lysolipid | [M-H] | 424.2484 | 0.10 [-0.27 - 0.48] | 5.84E-01 | -0.34 [-0.78 - 0.06] | 1.08E-01 |
| LPE (15:0), Peak 2 | Lysolipid | [M-H] | 438.2631 | 0.33 [-0.26 - 0.94] | 2.85E-01 | -0.61 [-1.35 - 0.08] | 8.96E-02 |
| LPE (16:0), Peak 1 | Lysolipid | [M-H] | 452.2788 | 0.14 [-0.58 - 0.87] | 7.01E-01 | -1.24 [-2.23 - -0.38] | 7.87E-03 |
| LPE (16:0), Peak 2 | Lysolipid | [M-H] | 452.2787 | 0.16 [-0.55 - 0.88] | 6.64E-01 | -0.51 [-1.39 - 0.26] | 2.00E-01 |
| LPE (16:1), Peak 2 | Lysolipid | [M-H] | 450.2629 | 0.26 [-0.11 - 0.65] | 1.70E-01 | 0.00 [-0.41 - 0.49] | 9.96E-01 |
| LPE (17:0), Peak 1 | Lysolipid | [M-H] | 466.2921 | 0.20 [-0.07 - 0.49] | 1.55E-01 | -0.07 [-0.37 - 0.23] | 6.62E-01 |
| LPE (17:0), Peak 2 | Lysolipid | [M-H] | 466.2939 | 0.15 [-0.31 - 0.62] | 5.30E-01 | -0.61 [-1.25 - -0.05] | 4.35E-02 |
| LPE (17:1), Peak 1 | Lysolipid | [M-H] | 464.2784 | 0.34 [-0.16 - 0.86] | 1.82E-01 | -0.23 [-0.75 - 0.27] | 3.66E-01 |
| LPE (17:1), Peak 2 | Lysolipid | [M-H] | 464.2784 | 0.45 [-0.07 - 1.00] | 9.41E-02 | -0.29 [-0.94 - 0.33] | 3.67E-01 |

| ID | class | adduct | m/z | irAE log(OR) | irAE pval | RECIST log(OR) | RECIST pval |
|---|---|---|---|---|---|---|---|
| LPE (18:0), Peak 1 | Lysolipid | [M-H] | 480.3077 | -0.04 [-0.65 - 0.58] | 9.09E-01 | -0.59 [-1.37 - 0.13] | 1.16E-01 |
| LPE (18:0), Peak 2 | Lysolipid | [M-H] | 480.31 | -0.04 [-0.83 - 0.75] | 9.26E-01 | -1.09 [-2.10 - -0.19] | 2.32E-02 |
| LPE (18:1), Peak 1 | Lysolipid | [M-H] | 478.2943 | 0.10 [-0.42 - 0.63] | 7.03E-01 | -0.38 [-1.02 - 0.23] | 2.34E-01 |
| LPE (18:1), Peak 2 | Lysolipid | [M-H] | 478.2939 | 0.06 [-0.52 - 0.64] | 8.45E-01 | -0.10 [-0.75 - 0.59] | 7.63E-01 |
| LPE (18:2), Peak 1 | Lysolipid | [M-H] | 476.2781 | 0.07 [-0.41 - 0.56] | 7.71E-01 | -0.49 [-1.12 - 0.10] | 1.12E-01 |
| LPE (18:2), Peak 2 | Lysolipid | [M-H] | 476.2788 | 0.06 [-0.36 - 0.48] | 7.85E-01 | -0.11 [-0.59 - 0.39] | 6.09E-01 |
| LPE (18:3), Peak 1 | Lysolipid | [M-H] | 474.263 | 0.00 [-0.40 - 0.41] | 9.82E-01 | -0.37 [-0.85 - 0.09] | 1.19E-01 |
| LPE (18:3), Peak 2 | Lysolipid | [M-H] | 474.2628 | 0.03 [-0.33 - 0.40] | 8.67E-01 | -0.16 [-0.57 - 0.25] | 4.32E-01 |
| LPE (20:2), Peak 1 | Lysolipid | [M-H] | 504.31 | 0.03 [-0.48 - 0.55] | 9.07E-01 | -0.35 [-0.97 - 0.23] | 2.38E-01 |
| LPE (20:2), Peak 2 | Lysolipid | [M-H] | 504.3099 | 0.41 [-0.12 - 0.97] | 1.38E-01 | -0.29 [-0.91 - 0.32] | 3.57E-01 |
| LPE (20:3), Peak 1 | Lysolipid | [M-H] | 502.2947 | 0.27 [-0.27 - 0.85] | 3.28E-01 | -0.46 [-1.16 - 0.19] | 1.76E-01 |
| LPE (20:3), Peak 2 | Lysolipid | [M-H] | 502.2946 | 0.17 [-0.33 - 0.67] | 5.06E-01 | -0.21 [-0.81 - 0.38] | 4.74E-01 |
| LPE (20:4), Peak 1 | Lysolipid | [M-H] | 500.2789 | 0.83 [0.03 - 1.70] | 4.99E-02 | -0.12 [-1.03 - 0.82] | 8.02E-01 |
| LPE (20:4), Peak 2 | Lysolipid | [M-H] | 500.279 | 1.07 [0.11 - 2.13] | 3.58E-02 | 0.01 [-1.05 - 1.11] | 9.83E-01 |
| LPE (20:5), Peak 1 | Lysolipid | [M-H] | 498.2632 | 0.14 [-0.31 - 0.60] | 5.40E-01 | -0.49 [-1.13 - 0.08] | 1.07E-01 |
| LPE (20:5), Peak 2 | Lysolipid | [M-H] | 498.2632 | 0.19 [-0.29 - 0.69] | 4.34E-01 | -0.47 [-1.12 - 0.12] | 1.31E-01 |
| LPE (22:4), Peak 1 | Lysolipid | [M-H] | 528.3026 | 0.48 [-0.46 - 1.44] | 3.10E-01 | 0.03 [-1.02 - 1.05] | 9.59E-01 |
| LPE (22:4), Peak 2 | Lysolipid | [M-H] | 528.3026 | 0.37 [-0.83 - 1.66] | 5.52E-01 | -1.45 [-3.37 - 0.10] | 1.01E-01 |
| LPI (16:0) | Lysolipid | [M-H] | 571.292 | 0.70 [-0.36 - 1.76] | 1.96E-01 | 0.73 [-0.64 - 2.11] | 2.94E-01 |
| LPI (18:0) | Lysolipid | [M-H] | 599.319 | 0.51 [-0.24 - 1.27] | 1.81E-01 | 0.52 [-0.35 - 1.55] | 2.86E-01 |
| LPI (18:1) | Lysolipid | [M-H] | 597.3002 | 0.17 [-0.07 - 0.42] | 1.65E-01 | 0.05 [-0.20 - 0.31] | 6.78E-01 |
| LPI (18:2) | Lysolipid | [M-H] | 595.2843 | -0.08 [-0.12 - 0.41] | 7.63E-01 | 0.00 [-0.59 - 0.59] | 9.93E-01 |
| LPI (20:4) | Lysolipid | [M-H] | 619.2946 | 0.12 [-0.23 - 0.46] | 5.04E-01 | 0.21 [-0.20 - 0.62] | 3.18E-01 |
| LysoPAF (16:0) | Lysolipid | [M+Cl-] | 516.3226 | 0.04 [-0.95 - 1.04] | 9.29E-01 | -1.22 [-2.65 - 0.05] | 7.38E-02 |
| 11-keto-testosterone | Sterol | [M-H+Acetate] | 361.2029 | -0.08 [-0.23 - 0.06] | 2.55E-01 | 0.07 [-0.08 - 0.24] | 3.55E-01 |
| Aldosterone | Sterol | [M-H+Acetate] | 419.2101 | 0.03 [-0.08 - 0.14] | 5.90E-01 | 0.03 [-0.09 - 0.16] | 6.03E-01 |

### Supplementary Table 4

#### Sub-Cohort from FINRISK-2002

| | Control | Autoimmune | p-value |
|---|---|---|---|
| n | 1712 | 158 | -- |
| Male (%) | 651 (38) | 54 (34) | 0.34 |
| Age (Range) | 39 (24-72) | 53 (25-74) | 0.003 |
| BMI (Range) | 24 (16-48) | 27 (18-41) | 0.45 |

#### Supplementary Table 5

| Description | Category | LPC (18:2) Effect Size | Standard Error | P-Value | LPC (16:0) Effect Size | Standard Error | P-Value |
|---|---|---|---|---|---|---|---|
| Other cheeses | Dairy | ***0.50*** | ***0.09*** | ***2.94E-06*** | 0.15 | 0.10 | 3.61E-01 |
| Low-fat cheese | Dairy | -0.21 | 0.09 | 1.21E-01 | 0.01 | 0.10 | 9.67E-01 |
| Cultured milk or yoghurt | Dairy | 0.07 | 0.09 | 6.95E-01 | -0.02 | 0.10 | 9.57E-01 |
| Sweet cookies or pies | Dessert, Carbohydrates | ***0.56*** | ***0.09*** | ***1.37E-07*** | 0.13 | 0.10 | 4.85E-01 |
| DELICACIES | Dessert, Carbohydrates | ***0.40*** | ***0.09*** | ***3.52E-04*** | 0.10 | 0.10 | 5.94E-01 |
| Sugared soft drinks | Dessert, Carbohydrates | ***0.33*** | ***0.10*** | ***8.70E-03*** | 0.23 | 0.10 | 1.35E-01 |
| Dark wheat or mixed grain bread | Dessert, Carbohydrates | ***0.31*** | ***0.09*** | ***1.13E-02*** | -0.14 | 0.10 | 4.45E-01 |
| FIBER_RICH_BREAD | Dessert, Carbohydrates | ***0.31*** | ***0.09*** | ***1.13E-02*** | -0.01 | 0.10 | 9.61E-01 |
| Low-calorie soft drinks | Dessert, Carbohydrates | -0.25 | 0.10 | 9.53E-02 | 0.03 | 0.11 | 9.19E-01 |
| Macaroni, pasta or rice | Dessert, Carbohydrates | -0.20 | 0.10 | 1.60E-01 | -0.22 | 0.10 | 1.35E-01 |
| Pastries | Dessert, Carbohydrates | 0.20 | 0.10 | 1.64E-01 | 0.07 | 0.10 | 7.27E-01 |
| Chocolate | Dessert, Carbohydrates | 0.20 | 0.09 | 1.71E-01 | -0.12 | 0.10 | 5.00E-01 |
| Rye bread, rye crisp | Dessert, Carbohydrates | 0.18 | 0.10 | 2.61E-01 | 0.13 | 0.11 | 5.14E-01 |
| Candy | Dessert, Carbohydrates | 0.07 | 0.09 | 6.95E-01 | 0.06 | 0.10 | 7.72E-01 |
| Ice cream etc | Dessert, Carbohydrates | 0.06 | 0.10 | 7.52E-01 | -0.04 | 0.10 | 8.31E-01 |
| White bread | Dessert, Carbohydrates | 0.06 | 0.10 | 7.72E-01 | 0.08 | 0.10 | 7.01E-01 |
| Diabetic diet | Diet | ***-1.38*** | ***0.31*** | ***2.64E-04*** | ***-1.14*** | ***0.30*** | ***3.64E-03*** |
| Weight diet | Diet | ***-0.91*** | ***0.22*** | ***7.54E-04*** | -0.10 | 0.23 | 8.29E-01 |
| Meals | Diet | ***0.30*** | ***0.10*** | ***2.09E-02*** | 0.25 | 0.10 | 8.82E-02 |
| Healthiness of eating habits / range 1-5, greater number equals unhealthier diet | Diet | -0.16 | 0.10 | 3.65E-01 | 0.19 | 0.11 | 2.93E-01 |
| Low cholesterol diet | Diet | -0.26 | 0.17 | 3.66E-01 | 0.17 | 0.18 | 6.37E-01 |
| FIBER_TOTAL | Diet | 0.12 | 0.09 | 4.94E-01 | 0.16 | 0.10 | 3.43E-01 |
| Lactose-free diet | Diet | 0.14 | 0.16 | 6.65E-01 | 0.02 | 0.17 | 9.60E-01 |
| Store bought ready-to-eat meals | Diet | 0.06 | 0.10 | 7.52E-01 | -0.18 | 0.10 | 3.09E-01 |
| Glutein-free diet | Diet | -0.23 | 0.51 | 8.28E-01 | -0.65 | 0.52 | 5.00E-01 |
| Vegetarian | Diet | 0.04 | 0.31 | 9.60E-01 | -0.35 | 0.32 | 5.84E-01 |
| Fresh salad and vegetables | Fruits, Vegetables | ***0.29*** | ***0.09*** | ***2.10E-02*** | 0.09 | 0.10 | 6.50E-01 |
| Fruit and berry juices | Fruits, Vegetables | 0.19 | 0.09 | 1.71E-01 | ***0.27*** | ***0.10*** | ***4.22E-02*** |
| Fresh or frozen berries | Fruits, Vegetables | -0.17 | 0.09 | 2.31E-01 | 0.07 | 0.10 | 7.20E-01 |
| HEALTHY_VEGETABLES | Fruits, Vegetables | 0.14 | 0.09 | 3.89E-01 | 0.16 | 0.10 | 3.38E-01 |

| | | | | | | | |
|---|---|---|---|---|---|---|---|
| Cooked vegetables | Fruits, Vegetables | 0.09 | 0.09 | 6.20E-01 | 0.18 | 0.10 | 2.31E-01 |
| Cooked or mashed potatoes | Fruits, Vegetables | 0.09 | 0.10 | 6.29E-01 | 0.13 | 0.10 | 4.91E-01 |
| Fruits | Fruits, Vegetables | -0.07 | 0.09 | 7.27E-01 | -0.02 | 0.10 | 9.57E-01 |
| Vegetarian foods | Fruits, Vegetables | 0.02 | 0.09 | 9.41E-01 | 0.06 | 0.10 | 7.43E-01 |
| FRUITS_BERRIES | Fruits, Vegetables | 0.01 | 0.09 | 9.74E-01 | 0.20 | 0.10 | 1.73E-01 |
| CEREALS | Grains or Nuts | 0.20 | 0.09 | 1.53E-01 | 0.02 | 0.10 | 9.57E-01 |
| Nuts | Grains or Nuts | 0.25 | 0.14 | 2.79E-01 | -0.09 | 0.15 | 7.52E-01 |
| Herbs | Grains or Nuts | -0.13 | 0.10 | 4.55E-01 | 0.10 | 0.10 | 6.29E-01 |
| Seeds | Grains or Nuts | 0.08 | 0.14 | 7.85E-01 | 0.01 | 0.15 | 9.76E-01 |
| Muesli or cereal | Grains or Nuts | 0.03 | 0.10 | 9.31E-01 | -0.02 | 0.10 | 9.57E-01 |
| Spreads | Oils | ***0.52*** | ***0.10*** | ***2.66E-06*** | 0.14 | 0.10 | 4.55E-01 |
| Dressing | Oils | ***0.32*** | ***0.09*** | ***8.70E-03*** | 0.02 | 0.10 | 9.57E-01 |
| Salmon | Protein | ***-0.53*** | ***0.10*** | ***2.66E-06*** | ***-0.31*** | ***0.10*** | ***2.20E-02*** |
| Fish and fish dishes | Protein | ***-0.40*** | ***0.10*** | ***7.54E-04*** | -0.21 | 0.10 | 1.60E-01 |
| Sausages and wieners | Protein | ***0.31*** | ***0.09*** | ***1.13E-02*** | 0.20 | 0.10 | 1.79E-01 |
| Herring | Protein | ***-0.33*** | ***0.11*** | ***2.77E-02*** | -0.17 | 0.12 | 4.12E-01 |
| WHITE_MEAT | Protein | ***-0.28*** | ***0.09*** | ***2.77E-02*** | -0.18 | 0.10 | 2.46E-01 |
| Cold cuts (e.g. salami) | Protein | 0.23 | 0.09 | 8.72E-02 | 0.14 | 0.10 | 4.12E-01 |
| Other fish | Protein | -0.21 | 0.09 | 1.37E-01 | 0.00 | 0.10 | 9.90E-01 |
| Soy | Protein | 0.35 | 0.16 | 1.45E-01 | 0.35 | 0.17 | 1.78E-01 |
| RED_MEAT | Protein | 0.18 | 0.09 | 2.15E-01 | 0.16 | 0.10 | 2.97E-01 |
| PROCESSED_MEAT | Protein | 0.17 | 0.09 | 2.31E-01 | 0.13 | 0.10 | 4.14E-01 |
| Cold meat cuts (e.g. cooked ham) | Protein | -0.14 | 0.09 | 3.89E-01 | 0.02 | 0.10 | 9.57E-01 |
| Meat dishes | Protein | 0.09 | 0.10 | 6.51E-01 | 0.18 | 0.10 | 2.79E-01 |
| Poultry meat | Protein | -0.07 | 0.10 | 7.21E-01 | -0.08 | 0.10 | 7.08E-01 |
| Eggs | Protein | 0.06 | 0.09 | 7.32E-01 | 0.06 | 0.10 | 7.77E-01 |
| JUNK_FOOD | Unhealthy Foods | ***0.29*** | ***0.09*** | ***2.20E-02*** | 0.08 | 0.10 | 7.01E-01 |
| Fried potatoes or french fries | Unhealthy Foods | ***0.28*** | ***0.10*** | ***3.97E-02*** | 0.10 | 0.10 | 6.22E-01 |
| Pizza | Unhealthy Foods | 0.27 | 0.10 | 7.92E-02 | -0.10 | 0.11 | 6.49E-01 |
| Salty snacks | Unhealthy Foods | 0.27 | 0.12 | 1.30E-01 | 0.11 | 0.12 | 6.75E-01 |
| Fast food | Unhealthy Foods | 0.05 | 0.13 | 8.73E-01 | 0.04 | 0.13 | 9.32E-01 |
| Burgers | Unhealthy Foods | 0.03 | 0.12 | 9.41E-01 | -0.02 | 0.13 | 9.55E-01 |
